# Legacy neuropsychiatric benefit after semaglutide is linked to maximum achieved dose and independent of the maximum weight lost

**DOI:** 10.64898/2026.04.16.26351060

**Authors:** Karthik Murugadoss, A.J. Venkatakrishnan, Venky Soundararajan

**Author notes:** Correspondence: Venky Soundararajan,.

## Abstract

GLP-1 receptor agonists have reshaped obesity therapeutics, but their impact on neuropsychiatric outcomes remains poorly characterized. From 29 million patients in a large federated data platform across the USA, including 489,785 semaglutide treated patients, we conducted an observational study integrating longitudinal neuropsychiatric outcomes. From this population, we assembled a cohort of 63,215 patients with baseline neuropsychiatric conditions before treatment initiation and evaluated 24 incident neuropsychiatric outcomes. In propensity-matched comparator analyses, during the 2 year time-period from treatment initiation, semaglutide was associated with broadly lower neuropsychiatric event risk than metformin, SGLT2 inhibitors, and DPP-4 inhibitors. Within the semaglutide-treated cohort, higher attained dose during the first two years after the first prescription (“pre-landmark period”) was associated with significantly lower incidence during the following two years (“post-landmark period”) of diagnostic codes associated with substance-related disorders (P<0.001), mood disorders (P<0.001), anxiety- and stress-related disorders (P<0.001), CNS atrophies (P<0.001), neuromuscular disorders (P=0.013), eating/sleep/behavioral disorders (P=0.022), and personality/impulse-control disorders (P=0.028). Consistent with previous clinical trials, the post-landmark incidence of dementia or CNS degenerative diseases was similar between the high-dose and low-dose semaglutide cohorts (P=0.15). For most neuropsychiatric diagnoses, post-landmark incidence was strongly associated with the maximum attained semaglutide dose during the pre-landmark period, but incident cognitive symptoms and speech/language symptoms were more closely linked to the pre-landmark weight-loss magnitude (p<0.001 and p<0.003, respectively). Bulk and single-cell transcriptomic analyses demonstrated GLP1R expression in CNS tissues (hypothalamus, caudate, putamen, nucleus accumbens, cerebellum) and peripheral nerves. Age-associated heterogeneity in GLP1R expression was evident in several of these compartments including the caudate nucleus, suggesting dynamic changes in the availability of the neurobiological substrate for semaglutide response. Together, these data support a model in which semaglutide confers a sustained, dose-dependent, weight loss-independent benefit across multiple neuropsychiatric conditions via direct CNS target engagement. This observational study motivates prospective clinical studies and mechanistic analyses to clarify the impact of GLP-1 receptor agonists on human neuropsychiatric pathways and disease processes.

## Introduction

While GLP-1 receptor agonists were originally positioned to improve glycemic control in type 2 diabetes, there is growing evidence from randomized clinical trials that these therapies also provide benefits in obesity, cardiovascular disease, chronic kidney disease, obesity-related heart failure with preserved ejection fraction, metabolic dysfunction associated liver disease, and sleep apnea^1–9^. These successes raise a practical translational question: when clinical benefit differs across disease domains, age strata, or gender, which aspects of that heterogeneity are due to direct pharmacology and which are indirect and secondary to systemic weight loss?

The answer is unlikely to come from whole-body summary measures alone. Human and translational studies now appreciate GLP1R biology across hypothalamic homeostatic circuits, hindbrain interoceptive pathways, and reward-linked striatal systems, while peripheral tissues such as heart, vasculature, gut, and endocrine organs may add additional on-target or context-dependent effects^10–17^. At the same time, tissue-level GLP1R localization outside canonical pancreatic compartments remains sensitive to assay choice and bulk tissue composition, making direct translational inference challenging.

To help deconvolute complex pharmacology of GLP-1 receptor agonists, particularly cognitive and neuropsychiatric outcomes, this work leverages EHR from a federated network of 489,785 semaglutide-treated patients^18–20^. We assessed 24 neuropsychiatric outcomes in a two-year period following semaglutide exposure to determine which clinical signals appear more compatible with direct CNS-linked activity and which appear more compatible with broader metabolic improvements. We then aligned those clinical signatures with transcriptomic datasets spanning the majority of human tissue types to build mechanistic hypotheses for these non-canonical clinical effects of semaglutide.

## Results

### Semaglutide shows broad advantage over metformin across affective, substance-use, vascular, and several cognitive-neurological outcomes

From a cohort of 489,785 patients with at least one semaglutide prescription, a neuropsychiatric analysis cohort of 63,215 patients with baseline neurological conditions was assembled (See **Methods**). Of these, 56,859 could be propensity matched 1:1 to metformin-treated patients, 4,757 to DPP-4 inhibitor-treated patients, and 30,680 to SGLT-2 inhibitor-treated patients. Across 24 incident neuropsychiatric outcomes, we evaluated approximate 2-year event risk after treatment initiation.

Semaglutide was associated with lower 2-year event risk than metformin (**Figure 1A**) for dementia/degenerative CNS disease (0.95% versus 1.96%; P < 0.001), cognitive symptoms (3.1% versus 4.3%; P < 0.001), substance-related neuropsychiatric disorders (3.8% versus 6.1%; P < 0.001), psychotic disorders (0.31% versus 0.93%; P < 0.001), speech/language symptoms (0.63% versus 1.21%; P < 0.001), cerebrovascular disease (2.6% versus 3.7%; P < 0.001), polyneuropathies (3.5% versus 4.5%; P < 0.001), mood disorders (10.5% versus 13.2%; P <0.001), personality/impulse-control disorders (0.53% versus 1.06%; P < 0.001), perceptual symptoms (0.24% versus 0.60%; P < 0.001), and neurocognitive disorders (1.0% versus 1.7%; P < 0.001). By contrast, metformin was associated with a lower rate of incident neuromuscular disease (1.1% versus 0.7%; P < 0.001).

**Figure 1.**
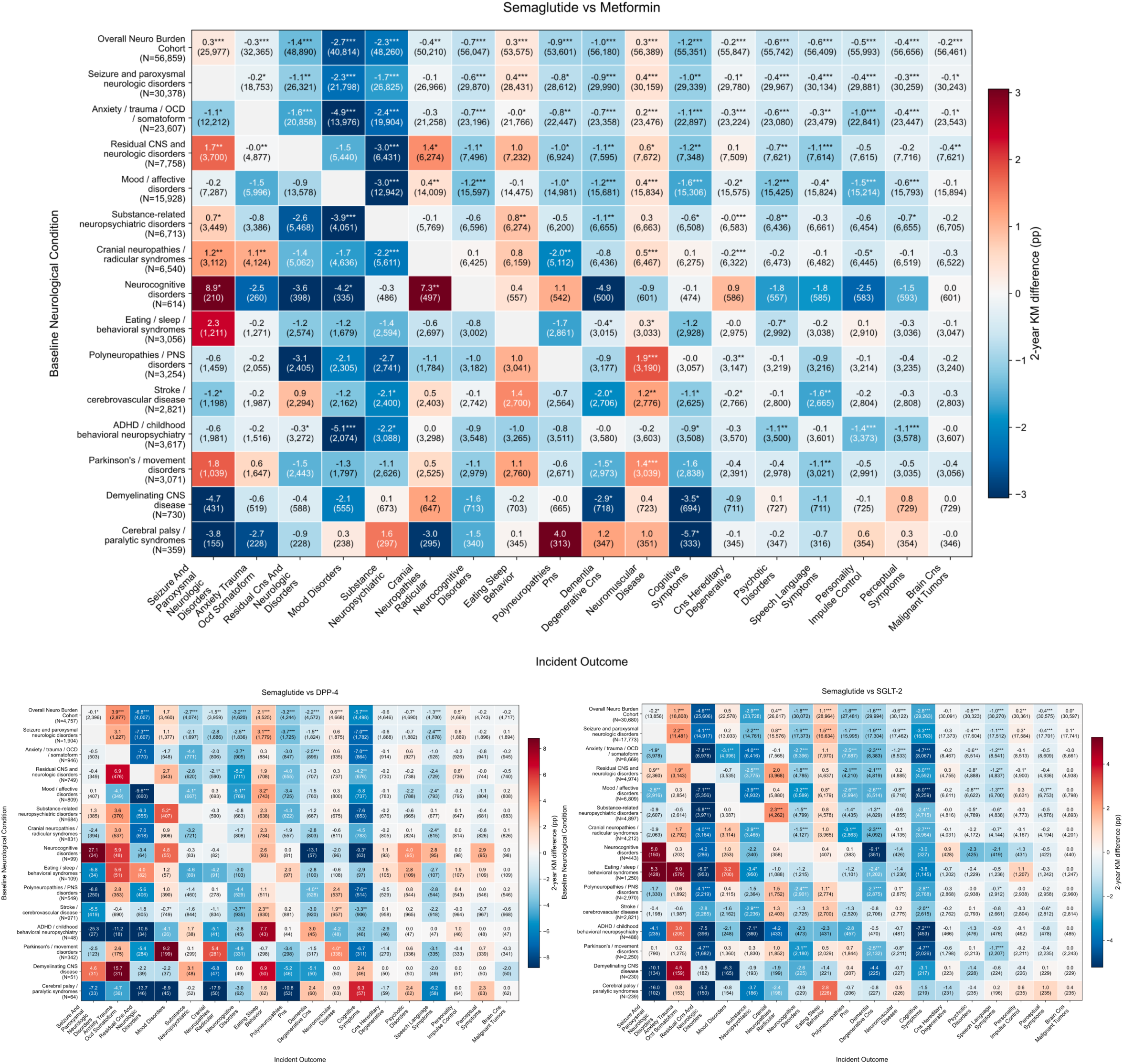
Semaglutide is broadly superior to metformin, DPP4 inhibitors and SGLT-2 inhibitors in the reduction of incident neurological and neuropsychiatric outcomes in the cohort with pre-recorded neurological/neuropsychiatric condition burden. Heat map showing 2-year Kaplan-Meier event-rate differences, in percentage points (pp), for semaglutide relative to metformin (*top),* DPP4 inhibitors (*bottom left)* and SGLT-2 inhibitors (*bottom right)* across baseline neurological condition (rows) and incident outcome (columns). Negative values *(blue*) indicate lower cumulative incident outcome with semaglutide, whereas positive values *(red*) indicate lower cumulative incidence with metformin. Cell annotations show the absolute 2-year difference in event probability, with the number of patients contributing to that comparison shown in parentheses; asterisks denote significance.

Subgroup-specific comparator analyses were then performed within individual baseline neurological-condition strata (see **Methods**). The largest semaglutide-favored effects were seen in patients with baseline neurocognitive disorders, where semaglutide was associated with lower subsequent mood disorders (13.7% versus 18.0%; P = 0.017). The strongest metformin-favored effects included lower rates of incident seizure and paroxysmal neurologic disorders among patients with baseline neurocognitive disorders (33.2% versus 24.3%; P = 0.015), as well as lower rates of incident neuromuscular disease among those with baseline polyneuropathy/peripheral nervous system disorders (3.0% versus 1.1%; P < 0.001) and Parkinson’s/movement disorders (2.2% versus 0.8%; P < 0.001). Broadly similar but more mixed patterns were observed in the corresponding comparisons with DPP-4 inhibitors (**Figure 1B**) and SGLT-2 inhibitors (**Figure 1C**), with semaglutide generally associated with generally more favorable associations across incident neuropsychiatric outcomes.

### Maximum attained semaglutide dose was associated with greater maximum weight loss through the landmark period

Maximum attained semaglutide dose during the first 2 years after treatment initiation, defined as the landmark period for subsequent analyses, was associated with greater maximum weight loss achieved within that same interval (**Figure 2**). Patients whose maximum dose remained at 0.25 mg or 0.5 mg showed the least weight reduction, whereas those reaching 1.7 mg, 2.0 mg, and especially 2.4 mg had progressively greater negative body-weight change distributions. The maximum achieved body-weight change increased by about 3.01% for each 1 mg increase in semaglutide dose. These findings indicate that dose attainment during the landmark period was strongly associated with the magnitude of achieved weight loss.

**Figure 2.**
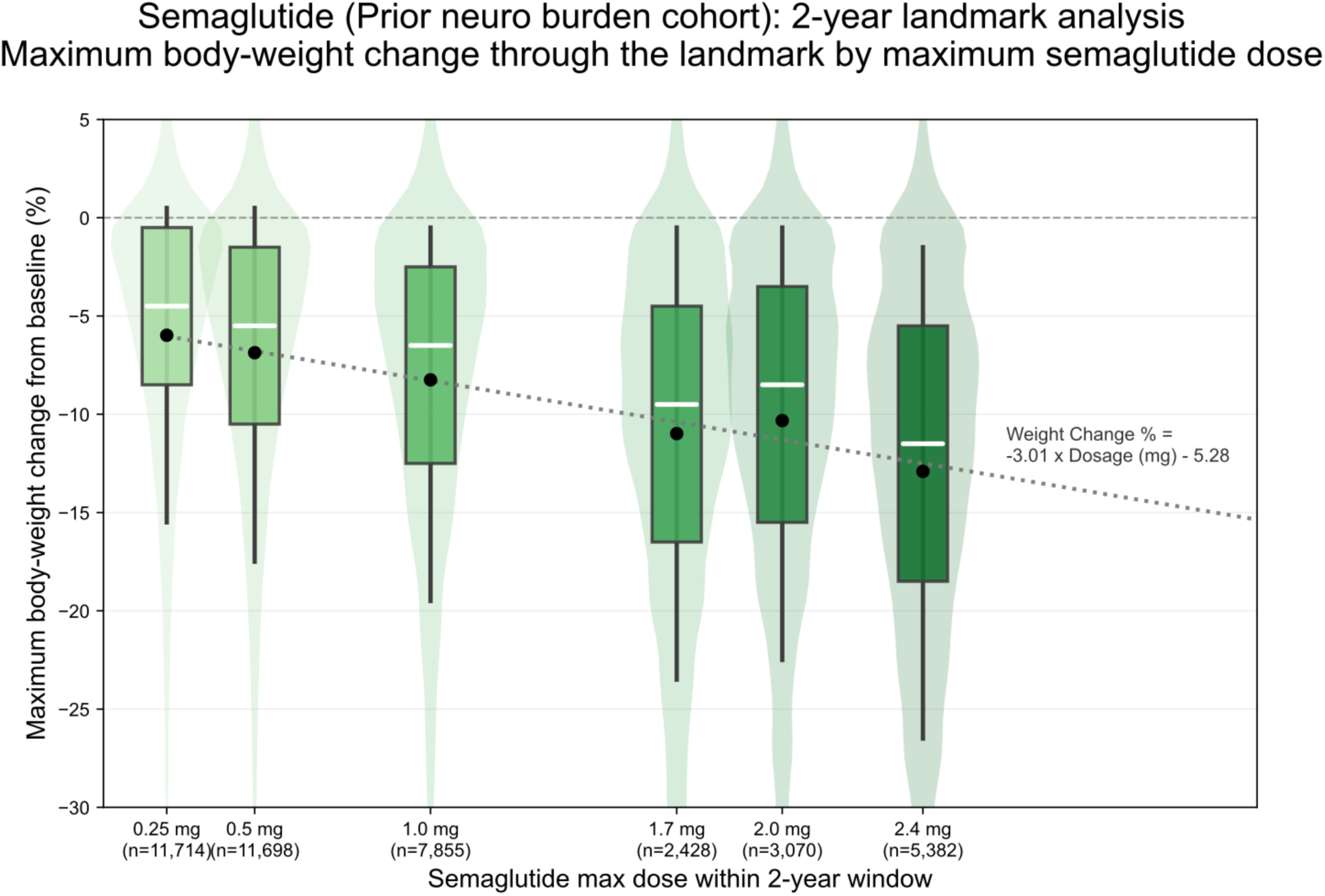
Maximum body weight change through the 2-year landmark by attained maximum semaglutide dose in this cohort with underlying neuropsychiatric/neurological conditions. Violin plots and embedded box plots show the distribution of maximum percent body weight change from baseline across semaglutide maximum dose groups attained within the 2-year landmark window (0.25 mg, 0.5 mg, 1.0 mg, 1.7 mg, 2.0 mg, and 2.4 mg). Black dots denote group means, white horizontal lines denote medians, boxes denote interquartile ranges, and whiskers denote distribution spread. The dotted regression line summarizes the dose-response trend, indicating progressively greater weight reduction with higher attained semaglutide dose. Sample sizes for each dose group are shown beneath the x-axis labels.

### Weight loss stratified analyses show little association with most neuropsychiatric outcomes

When patients were stratified by maximum achieved weight loss during the 0–2 year pre-landmark period, most post-landmark neuropsychiatric and neurological outcomes did not differ significantly across weight-loss categories (Figure 3, Figure S1). Baseline characteristics and semaglutide prescription patterns across weight-loss strata are summarized in Table 1. Global weight-loss-strata tests were non-significant for substance-related neuropsychiatric disorders (P = 0.15), mood/affective disorders (P = 0.96), and hereditary/systemic CNS atrophies (P = 0.99), with cumulative event curves showing no significant separation across the <5%, 5–10%, 10–15%, 15–20%, and ≥20% weight-loss groups. These findings suggest that, within this landmark framework, achieved weight loss was not strongly associated with the subsequent incidence of neuropsychiatric conditions.

**Figure 3.**
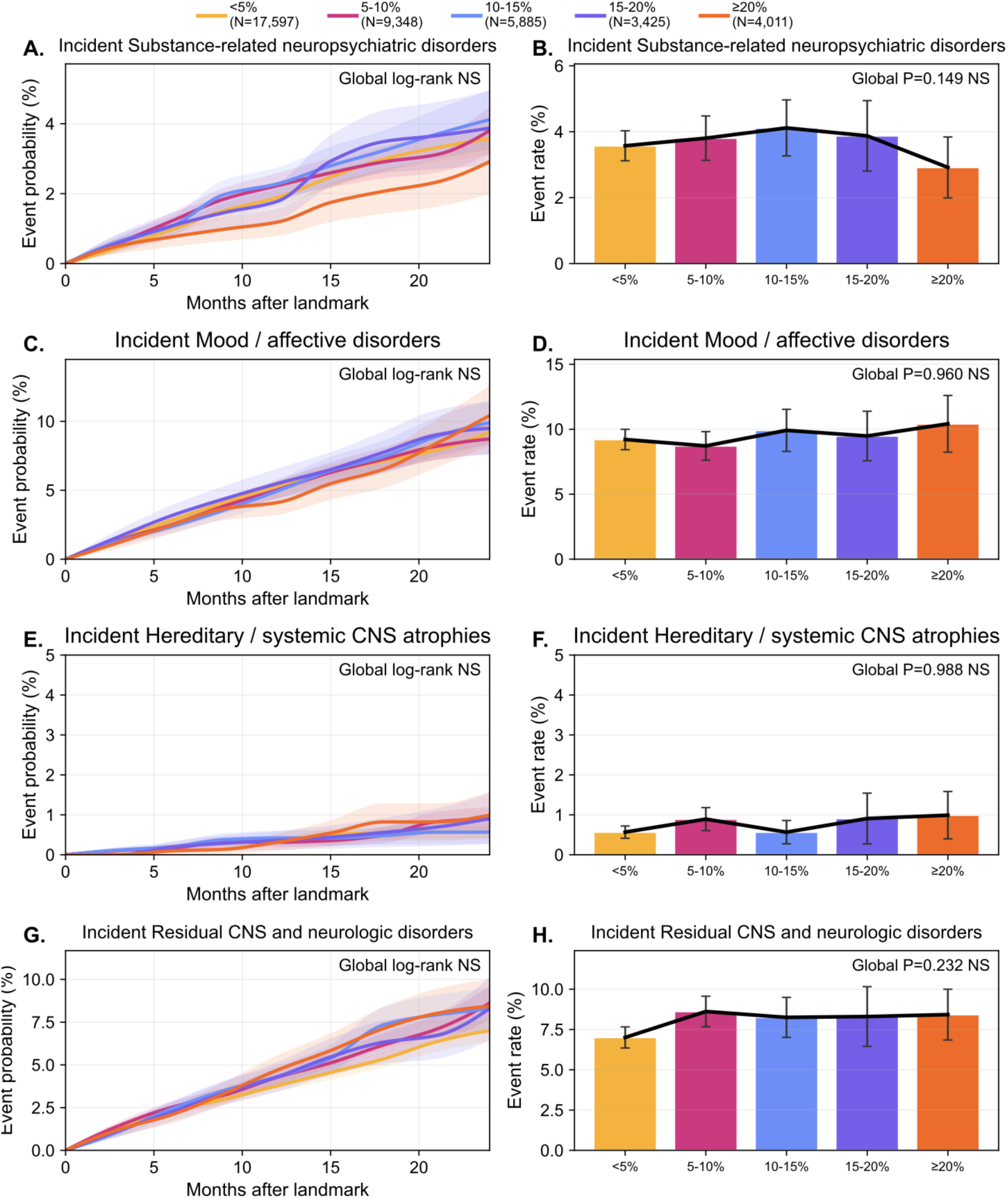
Post-landmark neurological and neuropsychiatric outcomes stratified by maximum achieved weight loss during semaglutide therapy in the pre-landmark period. Cumulative event curves and corresponding event-rate summaries are shown for substance-related neuropsychiatric disorders, mood/affective disorders, hereditary/systemic CNS atrophies, and residual CNS and neurologic disorders across five pre-landmark maximum weight-loss strata: <5%, 5–10%, 10–15%, 15–20%, and ≥20%. Panels on the left show post-landmark cumulative event probability from the time of landmark, and panels on the right show the corresponding event rates by weight-loss category. Global log-rank and event-rate P values are displayed within each panel.The neuropsychiatric and neurological outcomes shown were largely non-significant across categories.

**Table 1.** Baseline characteristics and pre-landmark (0-2 year) versus post-landmark (2-4 year) semaglutide prescription patterns by maximum attained weight loss.

| Weight-Loss Category | Number of Patients | Age, Mean (SD) | Female, % | Baseline BMI, Mean (SD) | Baseline Weight, kg, Mean (SD) | Baseline HbA1c, Mean (SD) | Baseline T2DM, n (%) | Max Dose >=1.7 mg, n (%) | Max Dose >=2.0 mg, n (%) | Max Dose >=2.4 mg, n (%) | Prescriptions, 0-2 Years Median [IQR] | Prescriptions, 2-4 Years (post-landmark) Median [IQR] | Treatment Duration, Days, Median [IQR] | Post-Landmark Follow-up, Days Median [IQR] |
| --- | --- | --- | --- | --- | --- | --- | --- | --- | --- | --- | --- | --- | --- | --- |
| All | 46362 | 53.4 (14.9) | 72.7 | 35.5 (6.1) | 103.1 (24.2) | 6.3 (1.5) | 7,926 (17.1%) | 13,254 (28.6%) | 10,809 (23.3%) | 7,713 (16.6%) | 2 [1, 5] | 0 [0, 0] | 33 [0, 276] | 176 [50, 419] |
| <5% | 20790 | 53.4 (15.1) | 69.8 | 35.3 (6.5) | 103.6 (25.8) | 6.3 (1.5) | 3,462 (16.7%) | 3,993 (19.2%) | 3,305 (15.9%) | 2,345 (11.3%) | 1 [1, 3] | 0 [0, 0] | 1 [0, 102] | 158 [42, 409] |
| 5-10% | 10724 | 54.2 (15.0) | 70.3 | 35.5 (6.0) | 103.0 (23.3) | 6.3 (1.4) | 1,911 (17.8%) | 3,019 (28.2%) | 2,451 (22.9%) | 1,705 (15.9%) | 2 [1, 5] | 0 [0, 0] | 49 [0, 267] | 172 [49, 417] |
| 10-15% | 6637 | 54.0 (14.7) | 74 | 35.6 (5.8) | 102.3 (22.5) | 6.4 (1.4) | 1,224 (18.4%) | 2,441 (36.8%) | 1,953 (29.4%) | 1,351 (20.4%) | 3 [1, 7] | 0 [0, 0] | 147 [0, 399] | 181 [57, 424] |
| 15-20% | 3799 | 52.8 (14.5) | 79 | 35.3 (5.7) | 100.7 (21.9) | 6.2 (1.3) | 651 (17.1%) | 1,622 (42.7%) | 1,304 (34.3%) | 928 (24.4%) | 4 [1, 9] | 0 [0, 0] | 216 [25, 474] | 206 [61, 438] |
| >=20% | 4,401 | 50.9 (14.5) | 85.3 | 36.5 (5.7) | 104.0 (22.8) | 6.2 (1.5) | 678 (15.4%) | 2,179 (49.4%) | 1,796 (40.7%) | 1,384 (31.4%) | 6 [2, 11] | 0 [0, 0] | 280 [39, 560] | 225 [79, 442] |

### Reduced rates of neuropsychiatric outcomes in patients achieving higher semaglutide doses

In the landmark analysis (Figure 4, Figure S2), higher attained semaglutide dose (≥1.7mg) between treatment initiation and the 2-year landmark was associated with lower post-landmark event rates than lower dose regimens (0.25-1 mg). Baseline characteristics and pre-versus post-landmark semaglutide prescription patterns across these dose strata are summarized in Table 2. Higher attained dose was associated with lower post-landmark event rates for substance-related neuropsychiatric disorders (RR 0.71, 95% CI 0.62-0.81; P < 0.001), mood/affective disorders (RR 0.82, 95% CI 0.75-0.89; P < 0.001, hereditary/systemic CNS atrophies (RR 0.59, 95% CI 0.45-0.78; P < 0.001), anxiety/trauma/OCD/somatoform disorders (RR 0.88, 95% CI 0.81-0.94; P < 0.001), neuromuscular junction or muscle disease (RR 0.72, 95% CI 0.55-0.93; P = 0.013), eating/sleep/behavioral syndromes (RR 0.87, 95% CI 0.76-0.99; P = 0.037), personality/impulse-control disorders (RR 0.68, 95% CI 0.48-0.96; P = 0.028), and neurologic signs and symptoms (RR 0.87, 95% CI 0.76-0.99; P = 0.037).

**Figure 4.**
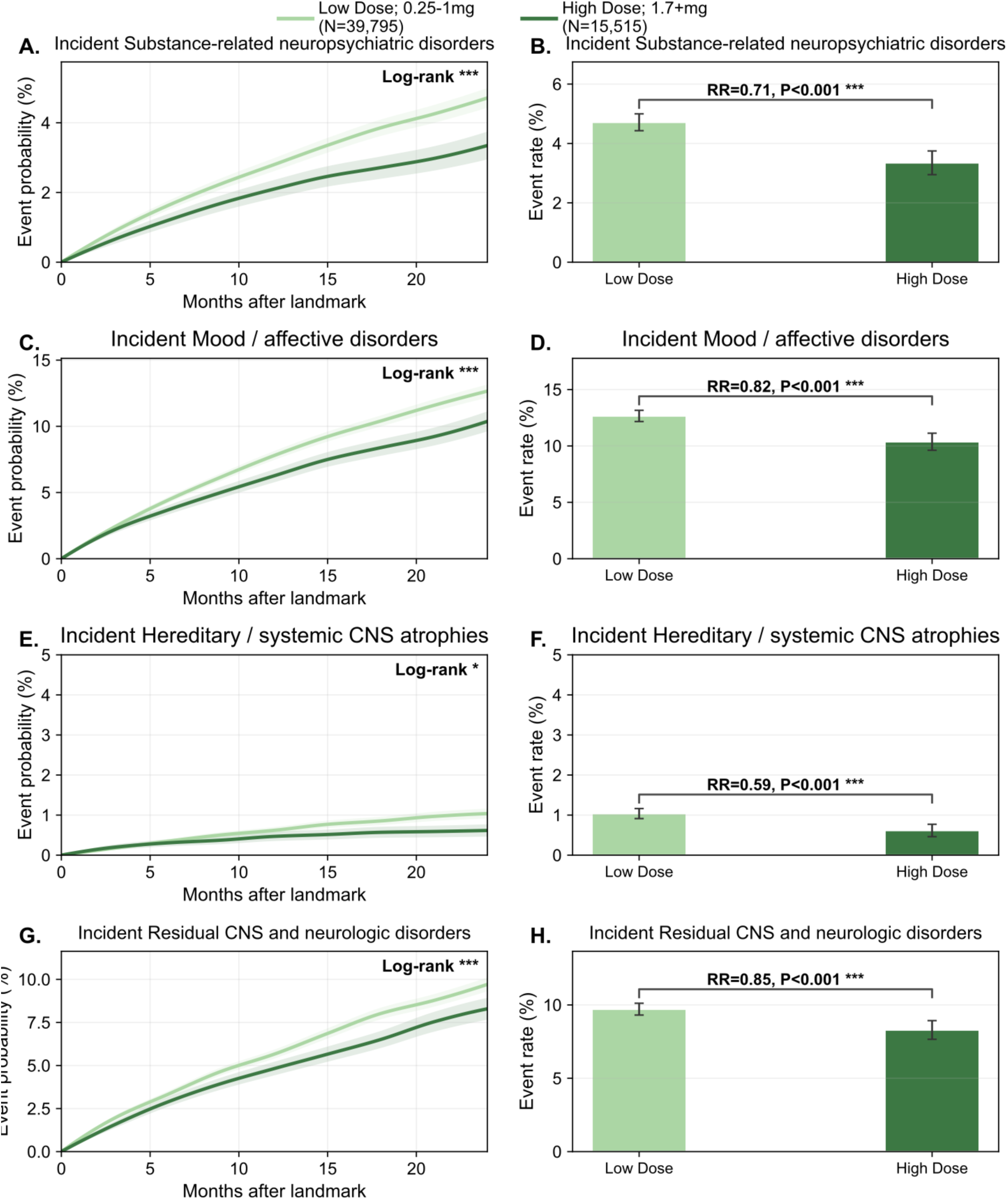
Post-landmark Neuropsychiatric outcomes by maximum attained semaglutide dose in the pre-landmark period. Cumulative event curves (left panels) and corresponding event-rate summaries (right panels) are shown for substance-related neuropsychiatric disorders, mood/affective disorders, hereditary/systemic CNS atrophies, and residual CNS and neurologic disorders after landmarking at 2 years from semaglutide initiation. Patients were grouped by maximum semaglutide dose attained during the pre-landmark window as low dose (0.25-1.0 mg; N = 39,795) or high dose (≥1.7 mg; N = 15,515). Across all outcomes shown, the high-dose group had lower post-landmark event rates than the low-dose group. Relative rate ratios and P values are displayed within the event-rate panels, and log-rank significance is shown within the cumulative-incidence panels.

**Table 2.**
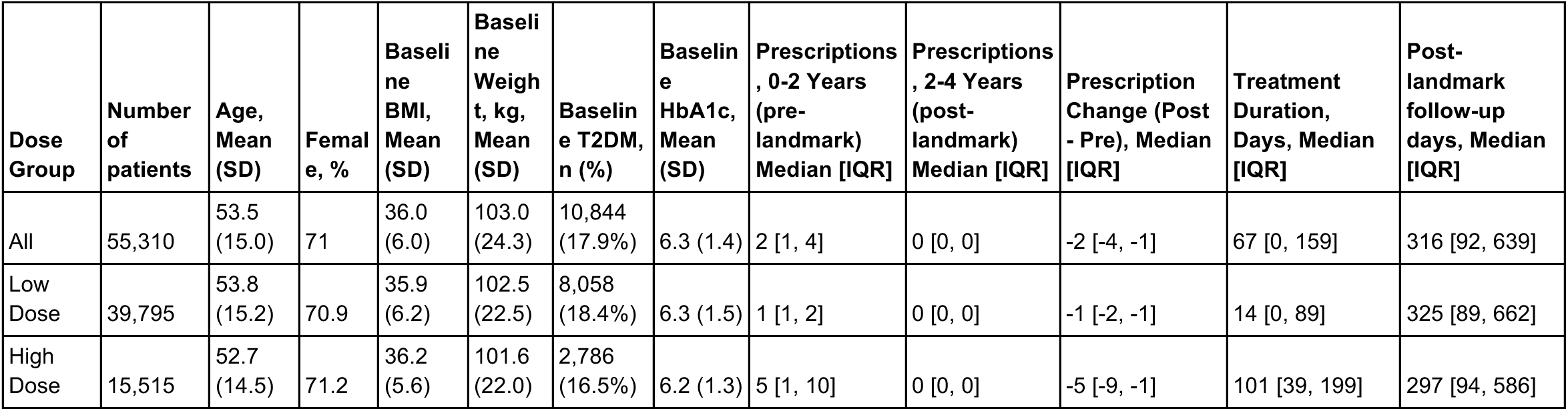
Baseline characteristics and pre-landmark (0-2 year) versus post-landmark (2-4 year) semaglutide prescription patterns by maximum attained dose.

| Dose Group | Number of patients | Age, Mean (SD) | Female, % | Baseline BMI, Mean (SD) | Baseline Weight, kg, Mean (SD) | Baseline T2DM, n (%) | Baseline HbA1c, Mean (SD) | Prescriptions, 0-2 Years (pre-landmark) Median [IQR] | Prescriptions, 2-4 Years (post-landmark) Median [IQR] | Prescription Change (Post - Pre), Median [IQR] | Treatment Duration, Days, Median [IQR] | Post-landmark follow-up days, Median [IQR] |
| --- | --- | --- | --- | --- | --- | --- | --- | --- | --- | --- | --- | --- |
| All | 55,310 | 53.5 (15.0) | 71 | 36.0 (6.0) | 103.0 (24.3) | 10,844 (17.9%) | 6.3 (1.4) | 2 [1, 4] | 0 [0, 0] | -2 [-4, -1] | 67 [0, 159] | 316 [92, 639] |
| Low Dose | 39,795 | 53.8 (15.2) | 70.9 | 35.9 (6.2) | 102.5 (22.5) | 8,058 (18.4%) | 6.3 (1.5) | 1 [1, 2] | 0 [0, 0] | -1 [-2, -1] | 14 [0, 89] | 325 [89, 662] |
| High Dose | 15,515 | 52.7 (14.5) | 71.2 | 36.2 (5.6) | 101.6 (22.0) | 2,786 (16.5%) | 6.2 (1.3) | 5 [1, 10] | 0 [0, 0] | -5 [-9, -1] | 101 [39, 199] | 297 [94, 586] |

### Weight-loss-dominant outcomes separate from the dose-dominant cluster

To distinguish dose-associated from weight-loss-associated patterns, post-landmark neurological and neuropsychiatric outcomes were next evaluated along two complementary axes: (a) the strength of association with attained dose and (b) the strength of association with achieved weight loss. Under this framework, substance-related neuropsychiatric disorders, mood/affective disorders, hereditary/systemic CNS atrophies, anxiety/trauma/OCD/somatoform disorders, and residual CNS and neurologic disorders clustered as predominantly dose-associated outcomes, whereas cognitive symptoms and speech/language symptoms showed stronger associations with weight-loss strata than with attained dose (**Figure 5**; **Table 3**). For cognitive symptoms, this weight-loss-associated pattern was particularly prominent (global P < 0.001), with approximate post-landmark event rates rising from 2.1% in the <5% weight-loss group to 5.5% in the 15–20% group and 4.7% in the ≥20% group (**Figure S3**). Speech/language symptoms showed a similar but smaller pattern (global P = 0.003). Together, these findings suggest that the favorable neuropsychiatric signals associated with semaglutide are not explained uniformly by greater weight reduction, but may instead reflect direct or CNS-adjacent pharmacology as well as frailty, disease burden, ascertainment bias, reverse causation, or symptom-linked treatment trajectories.

**Figure 5.**
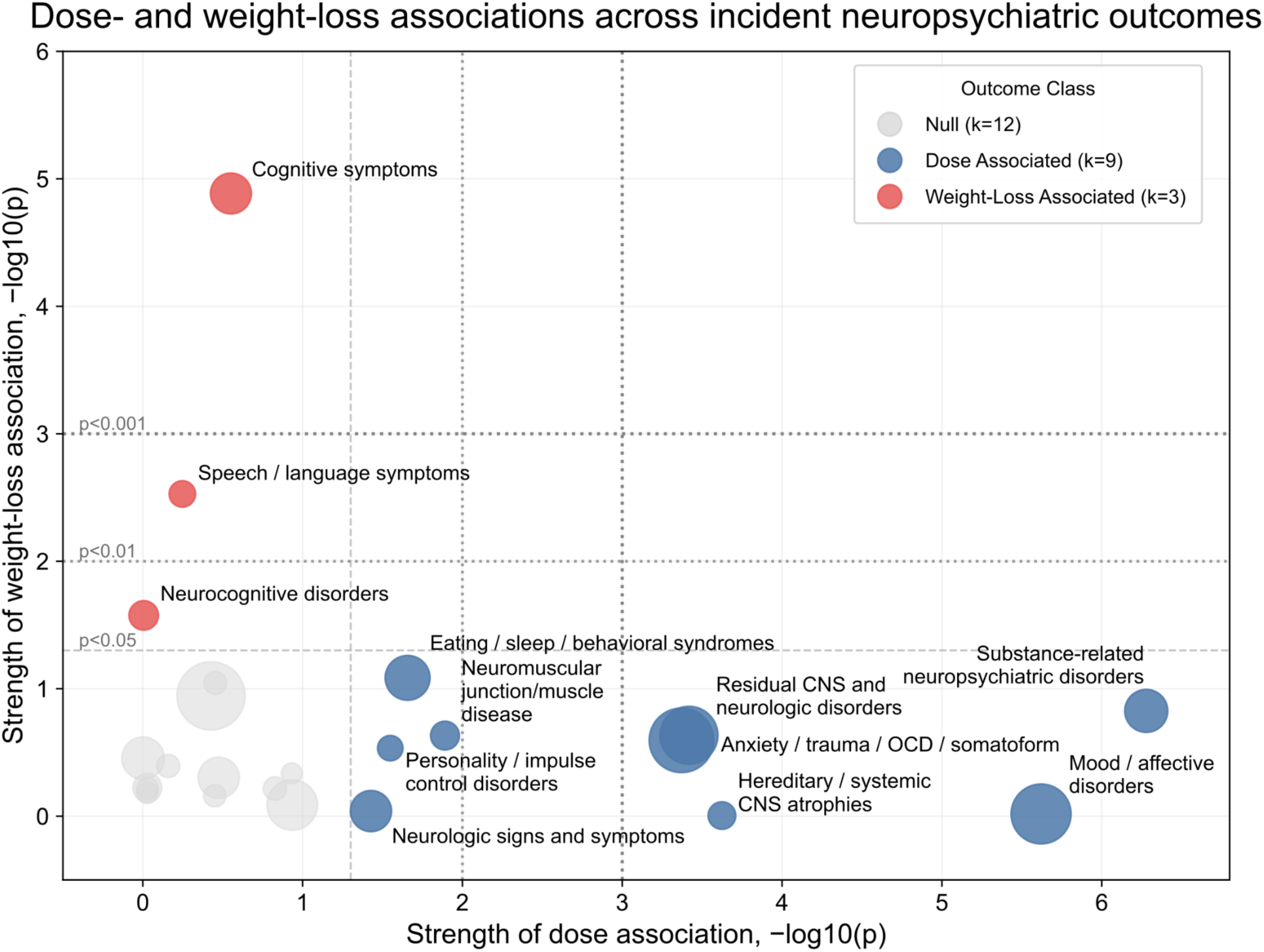
Dose- and weight-loss-associated incident neuropsychiatric outcomes among semaglutide-treated patients with baseline neurological disease burden. Each bubble represents one incident outcome from the 2-year landmark analyses in semaglutide-treated patients with prior neurological burden. The x axis shows the strength of association with maximum achieved semaglutide dose within the landmark window (−log10(P) for the low-versus high-dose comparison), and the y axis shows the strength of association with maximum achieved weight loss within the same landmark window (−log10(P) across weight-loss strata). Bubble area is proportional to the total number of incident events for that outcome. Outcomes are categorized as dose associated (blue), weight-loss associated (red) or null (grey); Higher values indicate stronger statistical evidence of association.

**Table 3.**
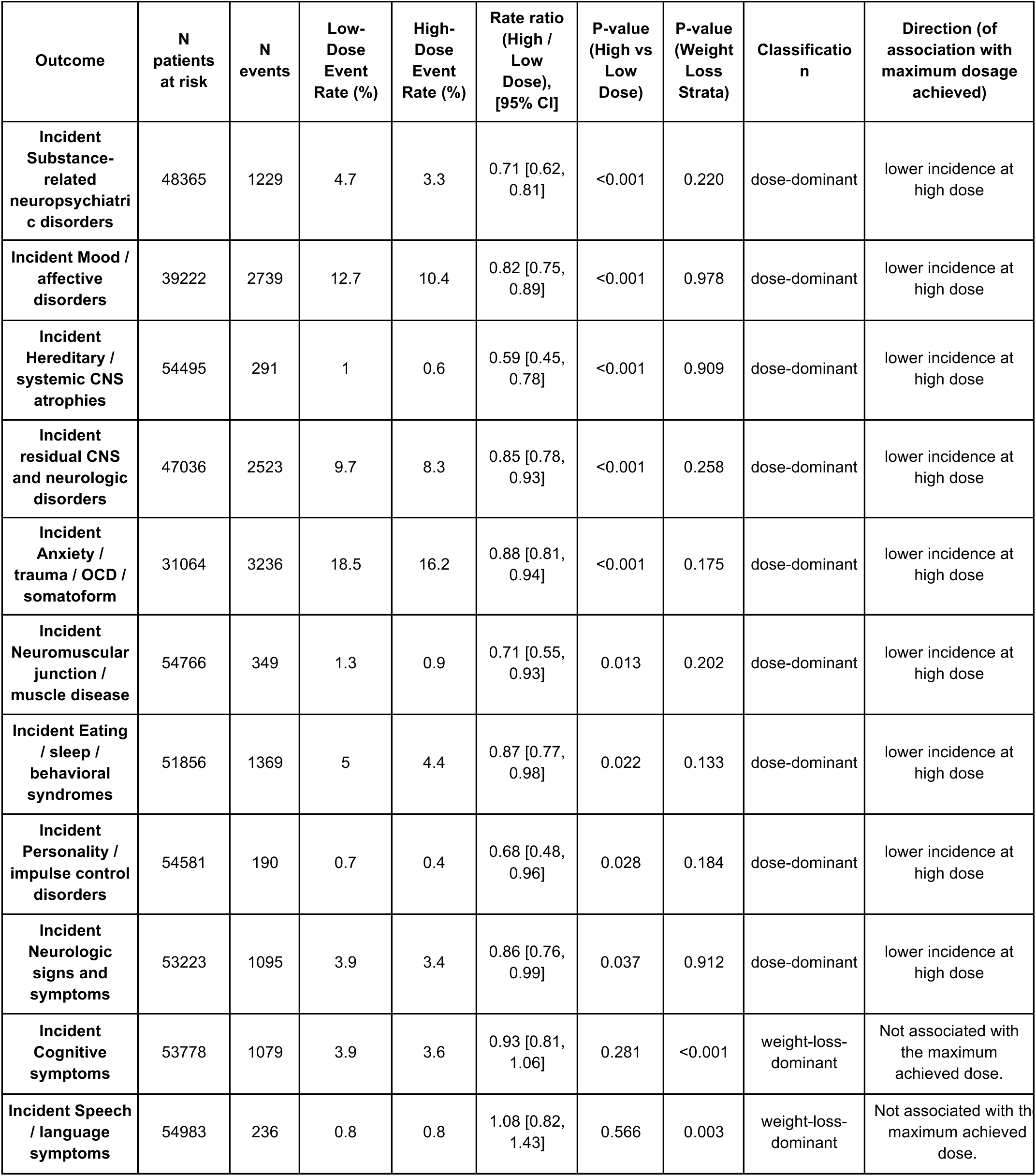
Cohort with underlying Neuropsychiatric conditions treated with Semaglutide in the pre-landmark period (0-2 years), summarizing the post-landmark (2-4 year) neuropsychiatric outcomes. Only outcomes showing significant dose- or weight-loss-associated differences are shown here; the remaining outcomes are provided in Table S2.

### A distributed human *GLP1R* atlas identifies candidate sites of expression in the central nervous system

As expected, bulk RNA-sequencing data across human tissues highlighted the pancreas as the highest *GLP1R*-expressing tissue, including endocrine and exocrine components (**Figure 6a, Table S3**). However, several non-pancreatic sites of *GLP1R* expression (median expression ≧ 1 TPM) were noted, including in the heart (atrial appendage), central nervous system (hypothalamus, caudate), and the peripheral nervous system (tibial nerve) (**Figure 6b**). Among the central nervous system (CNS) sites of possible expression, the highest expression was seen in the caudate nucleus of the basal ganglia (median 1.50 TPM, n=300) and hypothalamus (median 1.21 TPM, n=257), the latter being consistent with the well-established expression and critical function of GLP1R in hypothalamic neurons^21–25^. Of note, there was significant overlap in the *GLP1R* expression distributions of the hypothalamus and the other aforementioned CNS components, raising the possibility of additional neuronal circuitry that may contribute to the dose-associated clinical phenotypes observed in the present clinical dataset.

**Figure 6.**
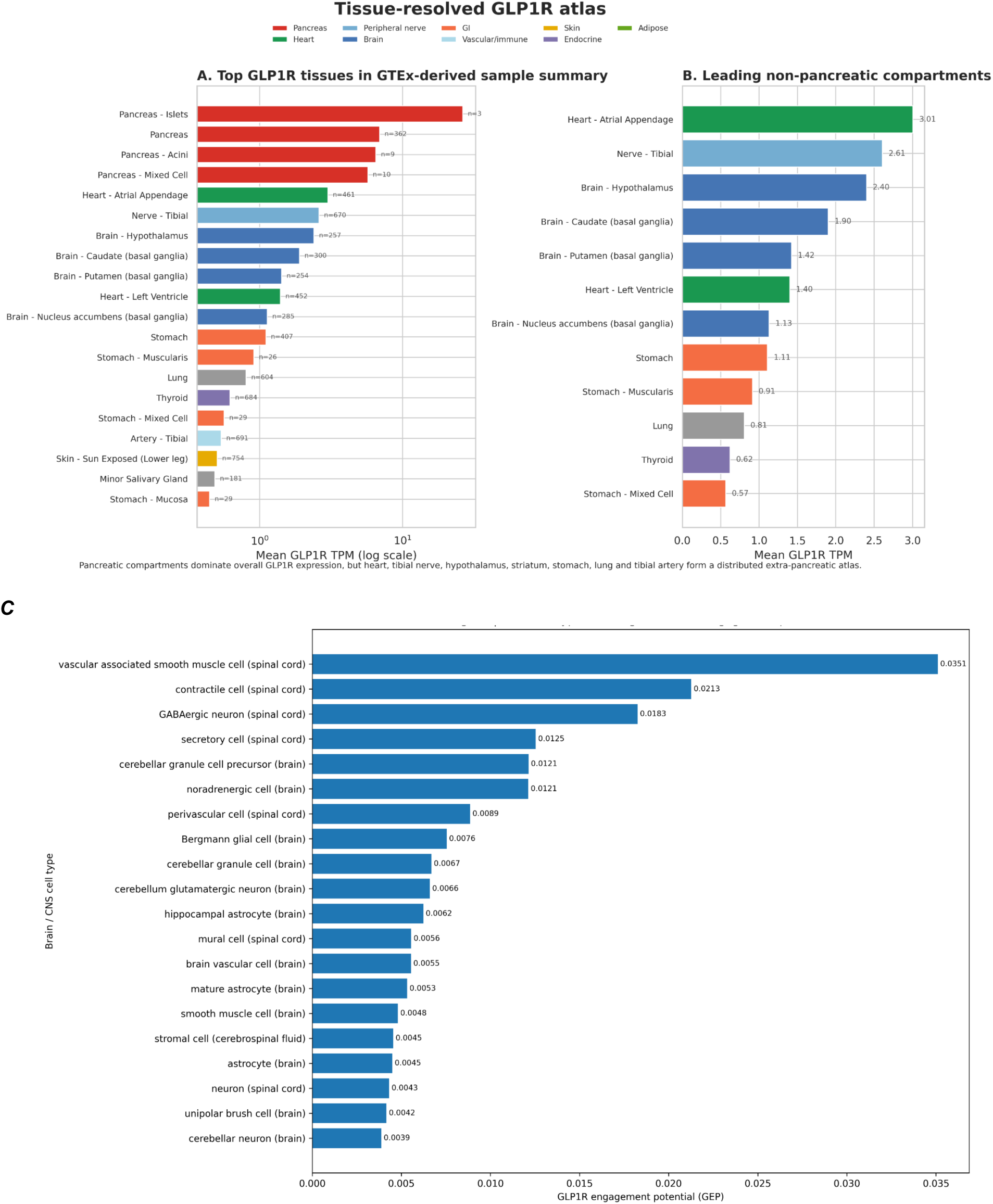
Tissue- and cell-resolved GLP1R expression. **(A)** Pancreatic compartments dominate global bulk RNA expression for GLP1R, but heart atrial appendage, tibial nerve, hypothalamus, basal ganglia, stomach, lung, thyroid, and tibial artery form a distributed extra-pancreatic landscape relevant to semaglutide and tirzepatide biology. **(B)** is the same as (A) without pancreas tissues. (**C**) shows significant GLP1R engagement potential (GEP) cells from the central nervous system (CNS) that are GLP1R+ from single cell RNA-seq.

### Single-cell *GLP1R* engagement potential identifies expression in neurovascular, astroglial, and neuronal CNS compartments

Single-cell RNA-seq analysis of the Central Nervous System (CNS) using cellxgene census obtained from the Chan Zuckerberg Initiative^26^ identified a restricted set of *GLP1R*-enriched cell types when ranked by *GLP1R engagement potential (GEP)*, defined as mean expression multiplied by the fraction of expressing cells (**Figure 6C**). The highest-ranking populations were vascular-associated smooth muscle cells in spinal cord (GEP = 0.0351), contractile cells in spinal cord (0.0213), and GABAergic neurons in spinal cord (0.0183), followed by secretory cells in spinal cord (0.0125), cerebellar granule cell precursors in brain (0.0121), noradrenergic cells in brain (0.0121), perivascular cells in spinal cord (0.0089), Bergmann glial cells (0.0076), cerebellar granule cells (0.0067), cerebellum glutamatergic neurons (0.0066), hippocampal astrocytes (0.0062), and brain vascular cells (0.0055). Together, these data indicate that at the cellular level, CNS *GLP1R* expression is distributed across neurovascular, astroglial, and neuronal cell compartments in multiple anatomic regions including the spinal cord, cerebellum, and hippocampus. Notably, hypothalamic cell populations did not emerge among the top-ranked GEP cell types, which may reflect either limited representation or annotation of hypothalamic datasets within the cellxgene census.

### Age-patterned GLP1R expression across CNS and CNS-related tissues highlights neurovascular and metabolic modifiers of clinical heterogeneity

Bulk RNA-sequencing data also suggested age-associated heterogeneity in *GLP1R* expression across nervous system and metabolically active tissues (**Figure 7A, Figure S4, Table S4**). Visceral adipose tissue showed a decreasing then increasing trajectory, with mean expression rising from 0.10 TPM in early adulthood to 0.20 by ages 70-79 years (P = 1.16 × 10⁻⁵; q = 1.16 × 10⁻⁵). For example, expression in the basal ganglia showed the largest dynamic range, declining from a mean expression of 4.17 TPM in patients aged 20–29 years to 1.23 TPM in patients aged 70–79 (P = 0.00124; q = 0.0109). Peripheral nerve expression also showed a complex nonmonotonic trajectory, rising from a mean of 1.82 TPM in patients aged 20–29 years to 2.99 TPM at 50–59 years before declining to 2.34 TPM at 70–79 years (P = 0.0036; q = 0.027). Together, these findings suggest tissue-specific dynamic changes in *GLP1R* expression over time.

**Figure 7.**
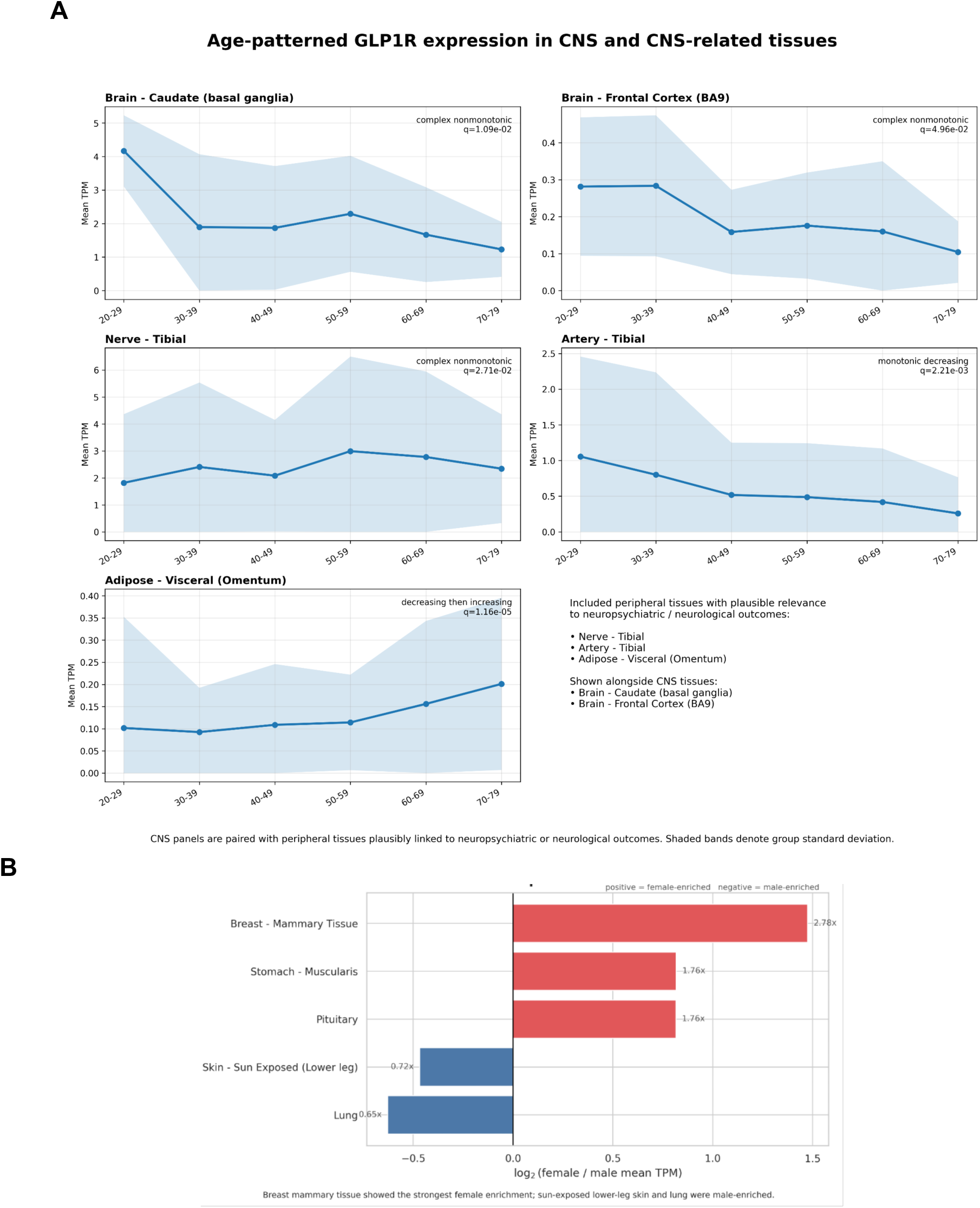
Demographic differences in GLP1R expression. **(A)** Mean GLP1R transcript abundance (TPM) across age brackets in selected tissues with plausible relevance to the observed neuropsychiatric and neurological outcome heterogeneity. CNS tissues shown are Brain - Caudate (basal ganglia) and Brain - Frontal Cortex (BA9). Peripheral tissues with potential neurobehavioral, neurovascular, or systemic relevance are Nerve - Tibial, Artery - Tibial, and Adipose - Visceral (Omentum). Artery - Tibial showed a monotonic decline with age, whereas Adipose - Visceral (Omentum) showed a decreasing-then-increasing trajectory; Brain - Caudate (basal ganglia), Brain - Frontal Cortex (BA9), and Nerve - Tibial showed more complex nonmonotonic patterns. Points denote age-bracket mean TPM values and shaded bands denote group standard deviation. q-values shown in each panel represent false-discovery-rate-adjusted significance across the full set of age-associated tissue tests. **(B)** Tissues with greater GLP1R expression in females (red; positive RR) versus greater GLP1R expression in males (blue; negative RR).

### Differential GLP1R expression by sex is most evident in mammary, gastric muscularis, pituitary, and lung tissues

Comparison of *GLP1R* expression in human bulk RNA-sequencing samples stratified by sex highlighted several tissues with differential expression (**Figure 7B, Table S5**). Among female-enriched tissues, mean GLP1R expression was higher in stomach muscularis (1.30 TPM in females versus 0.74 TPM in males; female-to-male fold change 1.76; p = 3.35 × 10⁻³; q = 3.97 × 10⁻²) and in pituitary (0.162 versus 0.092 TPM; fold change 1.76; p = 3.68 × 10⁻³; q = 3.97 × 10⁻²). GLP1R expression was also higher in breast mammary tissue (0.683 versus 0.246 TPM; fold change 2.78; p = 5.44 × 10⁻²⁵; q = 2.94 × 10⁻²³), although this difference is likely influenced by marked sex-specific differences in tissue composition. Given the limited expression of GLP1R in the pituitary and breast mammary tissue, bulk RNA-seq alone will not suffice to draw mechanistic insights and single cell RNA-seq as well as protein signaling data stratified by sex would be necessary to draw more mechanistic insights. Among male-enriched tissues, mean GLP1R expression was lower in females in lung (0.587 versus 0.907 TPM; fold change 0.65; p = 1.95 × 10⁻³; q = 3.51 × 10⁻²).

## Discussion

This study demonstrated that in the real-world clinical setting, the maximum attained semaglutide dose during the first two years after the first prescription were more closely associated with the subsequent incidence of neuropsychiatric outcomes than achieved weight loss. Specifically, higher attained dose was associated with lower subsequent incidence of conditions related to mood/affect, anxiety, substance use, neuromuscular function, and sleep disturbances. In contrast, most of these disease categories did not show significant separation across weight-loss strata. This dissociation argues against a simple model in which all favorable semaglutide-associated neurological signals are downstream consequences of greater body-weight reduction alone. The weight-loss independent neuropsychiatric associations subsequent to semaglutide exposure motivates future prospective studies including randomized trials and longer follow-ups post-treatment cessation. Importantly, this study also highlighted heterogeneity in neuropsychiatric outcomes during and after semaglutide exposure. For example, cognitive symptoms and speech/language symptoms did not share the same dose-linked pattern observed for mood, substance-related, anxiety-spectrum, and broader CNS outcomes; instead, they aligned more strongly with weight-loss strata, and cognitive-symptom event rates increased in the higher achieved-weight-loss groups. These findings should not be interpreted as evidence that semaglutide worsens cognition, and indeed prior studies have demonstrated a positive association between weight loss and improved cognitive function^27^. Rather, like the other aggregated landmark outputs presented here, these results likely reflect a mixture of biology and bias, including frailty enrichment, illness-related catabolism, differential surveillance, reverse causation, heterogeneous symptom coding, and the clinical reality that first recorded diagnosis often lags far behind biological onset. Weight-loss magnitude is challenging to consider as an exposure variable in retrospective analyses, as weight loss can be driven by both intentional (e.g., pharmacotherapy, lifestyle adjustments) and non-intentional (e.g., known or undiagnosed illnesses, declining reserve, malnutrition) factors. Future prospective studies will help to disentangle these confounders and more definitively characterize the impacts of semaglutide exposure and therapy-induced weight loss on neuropsychiatric phenotypes.

Existing literature evaluating the association between incretin therapies and neuropsychiatric outcomes has yielded mixed results. Recent clinical trials studying the impact of semaglutide on clinical progression in patients with early Alzheimer’s disease showed no benefit compared to placebo^28^. In line with this, our analysis did not find any significant difference between the high-dose and low-dose cohorts in the incident diagnosis rates of dementia or residual CNS and neurologic disorders during the post-landmark period. In a retrospective study of nearly 96,000 patients with depression and/or anxiety, semaglutide use was associated with significantly lower risk of worsening depression, worsening anxiety, and worsening substance use^29^. Conversely, in a propensity-matched retrospective analysis of over 300,000 obese patients without baseline neuropsychiatric conditions (bipolar disorder, schizophrenia, depression, anxiety, or suicidal ideation), use of GLP-1RA therapy was associated with significantly increased incidence of major depression, anxiety, and suicidal behavior^30^. Independent of the GLP-1 axis, multiple prior studies have found positive that intentional weight loss is associated with improvement in some neuropsychiatric outcomes including depression and cognitive function^27,31^. Mechanistically, there is strong evidence that incretin therapies function at least in part via CNS signaling pathways, with preclinical data demonstrating that GLP-1 receptor agonists activate hypothalamic neurons and establishing that CNS *GLP1R* expression is necessary to mediating their appetite suppression, weight loss, and counteracting of “molecular age”^23,32–34^. Whether broader CNS pathways that impact mood, anxiety, personality, and cognition are engaged by these medications warrants further exploration.

The tissue-resolved GLP1R expression analysis provides plausible mechanistic hypotheses for distinct dose- and weight loss-associated neuropsychiatric outcomes. Bulk transcriptomic data identified multiple potential CNS sites of *GLP1R* expression, including the hypothalamus, caudate, putamen, and nucleus accumbens. This distribution is compatible with a model in which mood, substance-related, and anxiety-spectrum outcomes may be modulated by GLP1R-linked circuits in hypothalamic and/or striatal reward centers. This model is supported by human neuroimaging studies and recent hypothalamic and hindbrain expression atlases that localize GLP1R to neuronal populations involved in satiety, aversion, interception, and neuroendocrine output^21–25^. Similarly, the observed GLP1R expression in peripheral nerve samples provides a plausible hypothesis for the semaglutide dose associations with diseases of the neuromuscular junction. The single-cell RNA-seq analyses further sharpen this picture by placing CNS *GLP1R* engagement potential in neuronal, astroglia, and neurovascular compartments. While these observations do not prove mechanism, they do suggest cellular and circuit level substrates for the clinical patterns observed here. The analysis of age- and sex-stratified bulk RNA-sequencing samples across tissue types highlight additional heterogeneity which could contribute to differences in treatment outcomes.

This study has several limitations. First, it is a retrospective observational study and is thereby vulnerable to multiple sources of bias and residual confounding. Second, disease incidence was defined by the first instance of a diagnostic code documented in the EHR, which inherently reflects diagnostic capture rather than biological onset and is prone to variability based on diagnostic coding practices. Third, the dose versus weight loss decomposition is informative but heuristic. There are challenges associated with retrospective analysis of both medication dosing (e.g., variable treatment adherence which is not well captured in the EHR) and weight loss (e.g., selection bias and classification of intentional versus unintentional weight loss). For example, by definition, the low-dose cohort includes patients who never received a prescription for a recommended maintenance dose of semaglutide. It is likely that some of these patients never actually initiated semaglutide or took very few doses before their dose escalation was discontinued. Without access to prescription claims data or detailed narratives of medication adherence, it is difficult to establish the degree of longitudinal medication exposure in this group, which likely varies widely between patients in the group. Fourth, the broad CNS and symptom-level groupings applied here are analytically useful but clinically heterogeneous. More fine-grained analyses are warranted. Fifth, the analysis indiscriminately considered patients who were prescribed any formulation of semaglutide, including Ozempic or Wegovy. While both of these medications have shown cardiometabolic improvements, there are important differences including their dose escalation regimens, approval timelines, typical clinical use cases, and underlying characteristics of the patient populations receiving these prescriptions. Regarding their dosing differences, it is important to note that the 1.0 mg weekly dose of Ozempic, which is considered a maintenance dose, is included in the “low dose” category in the current study, while the two maintenance doses of Wegovy (1.7 or 2.4 mg weekly) are both defined as “high dose.” Finally, the *GLP1R* expression analyses rely on bulk and single cell transcriptomic datasets. These modalities have inherent limitations relating to tissue composition, sampling, cell capturability, data sparsity, and possibly limited correlation with protein expression^35^.

In summary, this study reveals several intriguing neuropsychiatric associations with semaglutide therapy, most of which track more closely with the maximum attained dose than the magnitude of achieved weight loss. This motivates future clinical trials of GLP-1 receptor agonists with scopes that expand beyond weight loss, glycemic control, and cardiovascular outcomes, namely trials that prospectively evaluate psychiatric, sleep, substance-use, and neurologic end points. Ultimately, clinical decision support systems built to optimize incretin therapy may need to incorporate baseline neuropsychiatric burden, treatment persistence, and tissue-linked biology to identify which patients are most likely to experience dose-linked neurobehavioral benefit and which outcomes warrant close clinical follow-up.

## Methods

### Data Sources and Study Design

This study was conducted as a retrospective observational analysis using deidentified longitudinal electronic health record data from the nSights Federated EHR Network^20^. All analyses were performed on deidentified data. The study integrated semaglutide-associated neurological and neuropsychiatric outcome analyses from the federated EHR platform with GTEx-derived GLP1R expression data across tissues and sample-level metadata.

The clinical outcomes component comprised summary analyses spanning 24 neurological or neuropsychiatric outcomes, including counts, event-rate ratios, dose-group P values, and weight-loss-strata P values. The transcriptomic component comprised GTEx-based GLP1R sample-level TPM measurements together with tissue summaries and metadata-stratified association analyses^12^. The overall design was intended as a clinically oriented translational synthesis linking semaglutide-associated neuropsychiatric outcome patterns to tissue-resolved GLP1R biology.

### Study Cohorts and Exposure Definitions

The primary analytic cohort comprised 63,215 semaglutide-treated patients with at least one baseline neurological or neuropsychiatric condition before treatment initiation. Medication exposure was ascertained from prescription or medication event records using the recorded event timestamp. The index date was defined as the date of the first qualifying prescription for the cohort-defining medication or medication class. Follow-up data were available through 31 December 2025. To create incident-user comparator cohorts with minimal treatment contamination, separate mutually exclusive cohorts were defined for metformin, DPP-4 inhibitors, and SGLT2 inhibitors using the same index-date framework and exclusion logic. Other GLP-1 receptor agonist medications were treated as exclusion exposures when defining the semaglutide cohort.

### Demographic and Clinical Covariates

Demographic variables included age and sex. Age at index was obtained from the index medication record. Baseline BMI was defined as the BMI measurement closest to the index date, recorded from 365 days before through 1 day after the index date. Baseline weight for weight-loss analyses was defined separately as the weight measurement closest to the index date, recorded from 90 days before through 14 days after the index date. Baseline type 2 diabetes was defined by the presence of at least three encounters carrying a type 2 diabetes code on three distinct dates before the index date. Baseline neurological and neuropsychiatric disease burden was defined using prespecified ICD-9 and ICD-10 code prefixes across 31 clinically relevant domains, including mood/affective disorders, anxiety/trauma/OCD/somatoform disorders, substance-related neuropsychiatric disorders, cognitive symptoms, neurocognitive disorders, speech/language symptoms, neuromuscular disease, cerebrovascular disease, seizure, and paroxysmal neurologic disorders, and hereditary or degenerative CNS syndromes. The union of these domains defined an overall baseline neurological-burden cohort for the within-semaglutide landmark analyses, whereas individual baseline neurological subgroups were examined separately in comparative analyses. Complete diagnosis code definitions are provided in **Table S1**.

### Propensity-Matched Comparative Neuropsychiatric Analyses

For comparator analyses, three additional mutually exclusive incident-user cohorts were constructed for metformin, DPP-4 inhibitors, and SGLT2 inhibitors using the same index-date and exclusion logic described above. Within each baseline neurological or neuropsychiatric subgroup, we performed pairwise comparisons anchored on semaglutide as the primary exposure cohort. Separate analyses were conducted for semaglutide versus metformin, DPP-4 inhibitors, and SGLT2 inhibitors. Propensity scores were estimated with logistic regression using age at index, sex, baseline BMI, and baseline type 2 diabetes status. Patients were matched 1:1 without replacement using nearest-neighbor matching and a caliper of 0.2 on the propensity-score scale.

Follow-up for comparative outcome analyses began on the treatment initiation date. Patients were followed until the outcome of interest, death, or the last observed clinical record. For incident neurological and neuropsychiatric outcomes, patients with prevalent disease of the same type at baseline were excluded from that specific analysis. We summarized matched cohort size, event counts, person-time, and cumulative 2-year event estimates using Kaplan-Meier methods and log-rank P values.

### Neuropsychiatric Outcomes and Landmark Analyses

Within-semaglutide 2-year landmark analyses were conducted to assess associations of attained dose intensity and achieved weight loss with subsequent neurological and neuropsychiatric outcomes.

For dose-response analyses, each patient’s maximum achieved dose within 2 years after treatment initiation was paired with that patient’s maximum achieved change in total body weight during the same 2-year period (**Figure 2**). Dose categories corresponded to labeled dosing increments for injectable semaglutide formulations: 0.25 mg, 0.5 mg, 1.0 mg, 1.7 mg, 2.0 mg, and 2.4 mg or greater. Observed dose values were used when directly available in the medication record, and missing dose values were extracted from medication descriptions using a large language model-assisted dose-mapping workflow^36^.

For post-landmark outcome analyses, the landmark was defined as the 2-year timepoint after treatment initiation for both dose- and weight-loss-based analyses. For weight-loss-based analyses, maximum percent body-weight reduction was calculated from the lowest observed weight recorded from day 15 after index through the same 2-year landmark, and patients were categorized into prespecified weight-loss strata of <5%, 5-10%, 10-15%, 15-20%, 20-25%, and >=25% (**Figure 3**). For dose-based analyses, the maximum semaglutide dose reached between index and the 2-year landmark was classified as low dose (0.25-1.0 mg) or high dose (>=1.7 mg; **Figure 4**). For each incident outcome analysis, patients with evidence of that same condition before follow-up were excluded. Post-landmark follow-up extended for 24 months and continued until the outcome of interest, death, or the last observed clinical record, whichever occurred first.To compare dose-associated and weight-loss-associated patterns across outcomes, the corresponding dose- and weight-stratified P values were jointly visualized in a two-axis bubble plot (**Figure 5**).

### GTEx-derived GLP1R expression

The GTEx-derived GLP1R atlas comprised 18,576 sample-level TPM measurements across 64 tissues, together with tissue-level summary statistics and metadata-stratified association outputs (**Figure 6A-B**). Tissue-level ranking was based on mean TPM values from the tissue summary table. Age- and sex-associated expression patterns were defined according to the reported q values, with significance assessed at q < 0.05 (**Figure 7**). Age trajectories were interpreted using the supplied age-pattern classifications, including monotonic decreasing, increasing then decreasing, decreasing then increasing, and other nonmonotonic patterns (**Figure S4**). Sex effects were summarized according to the direction of the mean female-versus-male difference. Metadata associations were used primarily as interpretive and quality-context variables rather than as primary biological endpoints.

### Statistical Analysis

Continuous variables were summarized as means with standard deviations or medians with interquartile ranges, as appropriate, and categorical variables were summarized as counts and percentages. In the comparative neuropsychiatric analyses, between-cohort outcome differences were summarized using Kaplan-Meier methods, with significance assessed by log-rank testing; comparative heatmaps displayed absolute differences in approximate 2-year Kaplan-Meier event risk. In the semaglutide weight-loss landmark analyses, cumulative event distributions and corresponding event-rate comparisons across weight-loss strata were assessed using a global log-rank test. In the dose landmark analyses, cumulative event distributions were compared between low-dose and high-dose groups using log-rank tests, whereas post-landmark event-rate comparisons were summarized as relative risks with 95% confidence intervals and Wald-type chi-square P values derived from log risk-ratio estimates. Bubble-plot visualizations displayed dose-and weight-loss-associated evidence as -log10(P) values derived from these corresponding dose and weight analyses.

For the GTEx analyses, tissue-level and sample-level summary statistics were analyzed descriptively and visualized directly from the available TPM, P-value, and q-value outputs. For plotting purposes, P values reported as zero in summary exports were truncated to 1 × 10−6 to enable log-scale visualization. Because the clinical outputs were analyzed in aggregated form and the molecular outputs reflected summary-level bulk-tissue statistics, mechanistic interpretations were treated as hypothesis generating rather than causal, and no attempt was made to infer mediation or patient-level counterfactual effects.

### Data Source

This study analyzed de-identified EHR data from academic medical centers in the United States via the nference nSights Analytics Platform. Prior to analysis, all data underwent expert determination de-identification satisfying HIPAA Privacy Rule requirements (45 CFR §164.514(b)(1)), employing a multi-layered transformation approach for both structured data (cryptographic hashing of identifiers, date-shifting, geographic truncation) and unstructured clinical text (ensemble deep learning and rule-based methods with >99% recall for personally identifiable information detection)^37^. nference established secure data environments within each participating center, housing these de-identified patient data governed by expert determination. These de-identified data environments were specifically designed to enable data access and analysis without requiring Institutional Review Board oversight, approval, or exemption confirmation. Accordingly, informed consent and IRB review were not required for this study.

### Data Availability

This study involves the analysis of de-identified Electronic Health Record (EHR) data via the nference nSights Federated Clinical Analytics Platform (nSights). Data shown and reported in this manuscript were extracted from this environment using an established protocol for data extraction, aimed at preserving patient privacy. The data has been de-identified pursuant to an expert determination in accordance with the HIPAA Privacy Rule. Any data beyond what is reported in the manuscript, including but not limited to the raw EHR data, cannot be shared or released due to the parameters of the expert determination to maintain the data de-identification. The corresponding author should be contacted for additional details regarding nSights.

### De-identification and HIPAA compliance certification

Prior to analysis, all EHR data were de-identified under an expert determination consistent with the Health Insurance Portability and Accountability Act (HIPAA) Privacy Rule (45 CFR §164.514(b)(1)). The de-identification methodology employed a multi-layered transformation approach to both structured and unstructured data fields^37^. In structured data, direct identifiers including patient names and precise geographic locations were excluded entirely, while indirect identifiers underwent specific transformations: patient identifiers, medical record numbers, and accession numbers were replaced with one-way cryptographic hashes using confidential salts to preserve linkage across patient encounters; all dates were shifted backward by patient-specific random offsets (1–31 days) to preserve temporal relationships while obscuring exact event timing; the ZIP codes were truncated to two-digit state-level resolution; and continuous variables including age, height, weight, and body mass index were thresholded to prevent identification of extreme values (for example, ages ≥89 years transformed to ‘89+’ and BMI >40 transformed to ‘40+’). In unstructured clinical text, an ensemble de-identification system that combines attention-based deep learning models with rule-based methods achieved an estimated >99% recall for personally identifiable information (PII) detection, with detected identifiers replaced by plausible fictional surrogates^37^.

### Data Harmonization

To address heterogeneity in EHR data, we harmonized clinical variables including medications, anthropometric measurements, and diagnoses to standardized concepts. For medications, we first constructed a standardized drug concept database combining the nSights knowledge graph with RXNorm (https://www.nlm.nih.gov/research/umls/rxnorm/index.html) hierarchies to capture ingredient, brand, and dose-specific information. EHR medication records were matched using a hierarchical approach prioritizing RXNorm codes when available, followed by ingredient-level matching, and finally natural language processing and pattern matching on free-text medication orders when structured codes were absent. For anthropometric measurements (height, weight, BMI), we created a unified vocabulary from SNOMED (https://www.snomed.org/, https://athena.ohdsi.org) and LOINC (https://loinc.org/) terminologies and matched EHR measurement descriptions using standardized text matching algorithms with abbreviation expansion and synonym resolution; ambiguous mappings were resolved using OpenAI GPT-4o^38^ (https://platform.openai.com/docs/models/gpt-4o) with summary statistics as context, followed by manual verification. For diagnoses, we developed a hierarchical disease concept database from the nSights knowledge graph and matched EHR diagnosis descriptions and codes by identifying the most specific common child concept in the hierarchy. This approach enabled consistent identification of clinical entities while preserving granularity where available.

## Acknowledgements

We thank the nference engineering team for the development of the nSights federated AI platform, and Patrick Lenehan and Christopher Gregg for critical review and manuscript feedback.

## Supplementary Material

**Figure S1.**
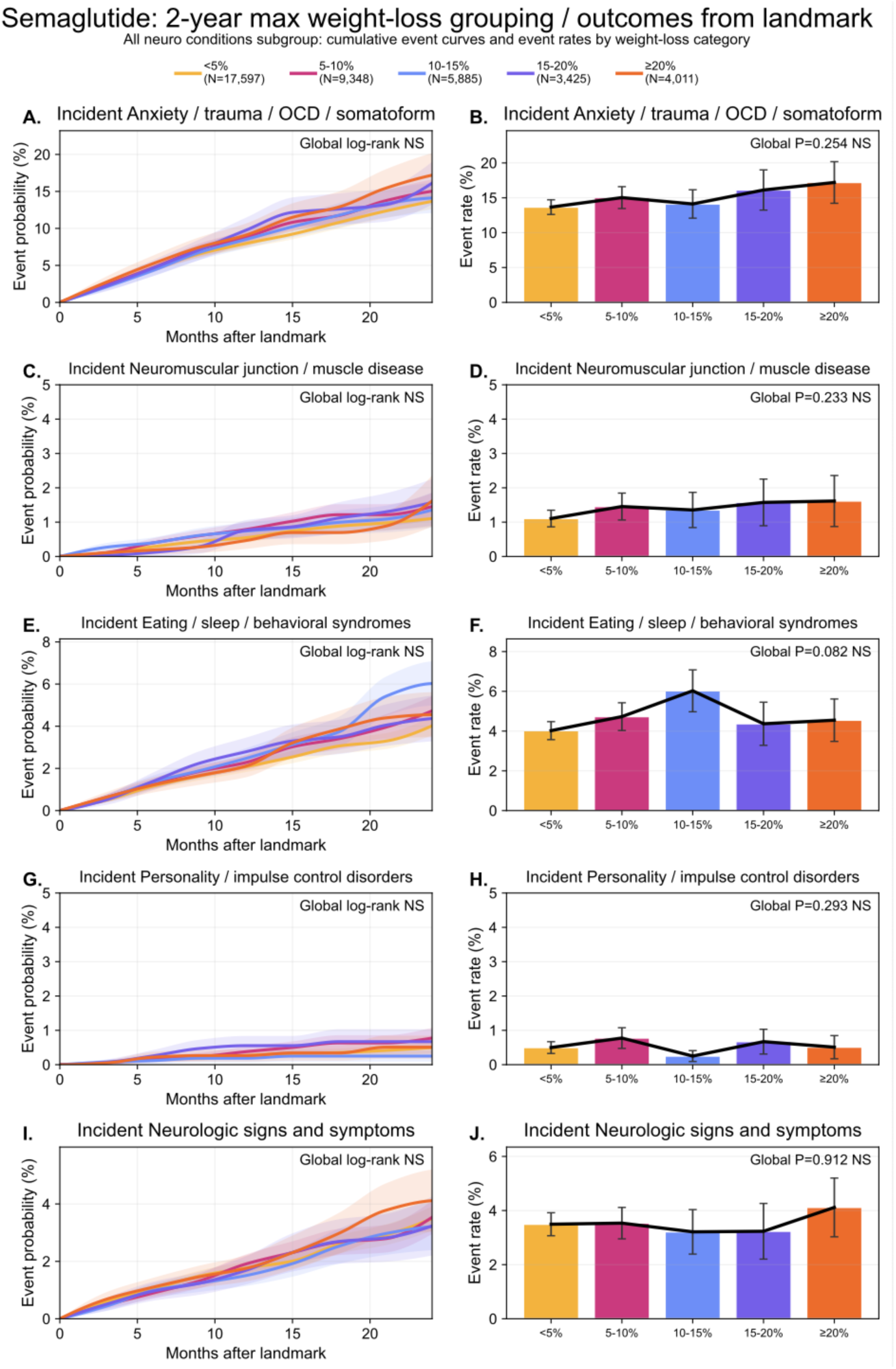
Post-landmark neurological and neuropsychiatric outcomes stratified by maximum achieved weight loss during the 0–2 year landmark period. Cumulative event curves (left panels) and corresponding event-rate summaries (right panels) are shown for anxiety/trauma/OCD/somatoform disorders, neuromuscular junction or muscle disease, eating/sleep/behavioral syndromes, personality/impulse-control disorders, and neurologic signs and symptoms across five categories of maximum body-weight change achieved before landmark: <5%, 5–10%, 10–15%, 15–20%, and ≥20%. Global log-rank statistics for the cumulative-incidence curves and global P values for the event-rate comparisons are displayed within each panel. None of the outcomes shown reached statistical significance across weight-loss strata, although eating/sleep/behavioral syndromes showed a numerically more variable pattern with a peak in the 10–15% weight-loss group. Overall, these analyses indicate limited evidence that the delayed post-landmark incidence of these domains was strongly determined by the magnitude of pre-landmark weight loss alone.

**Figure S2.**
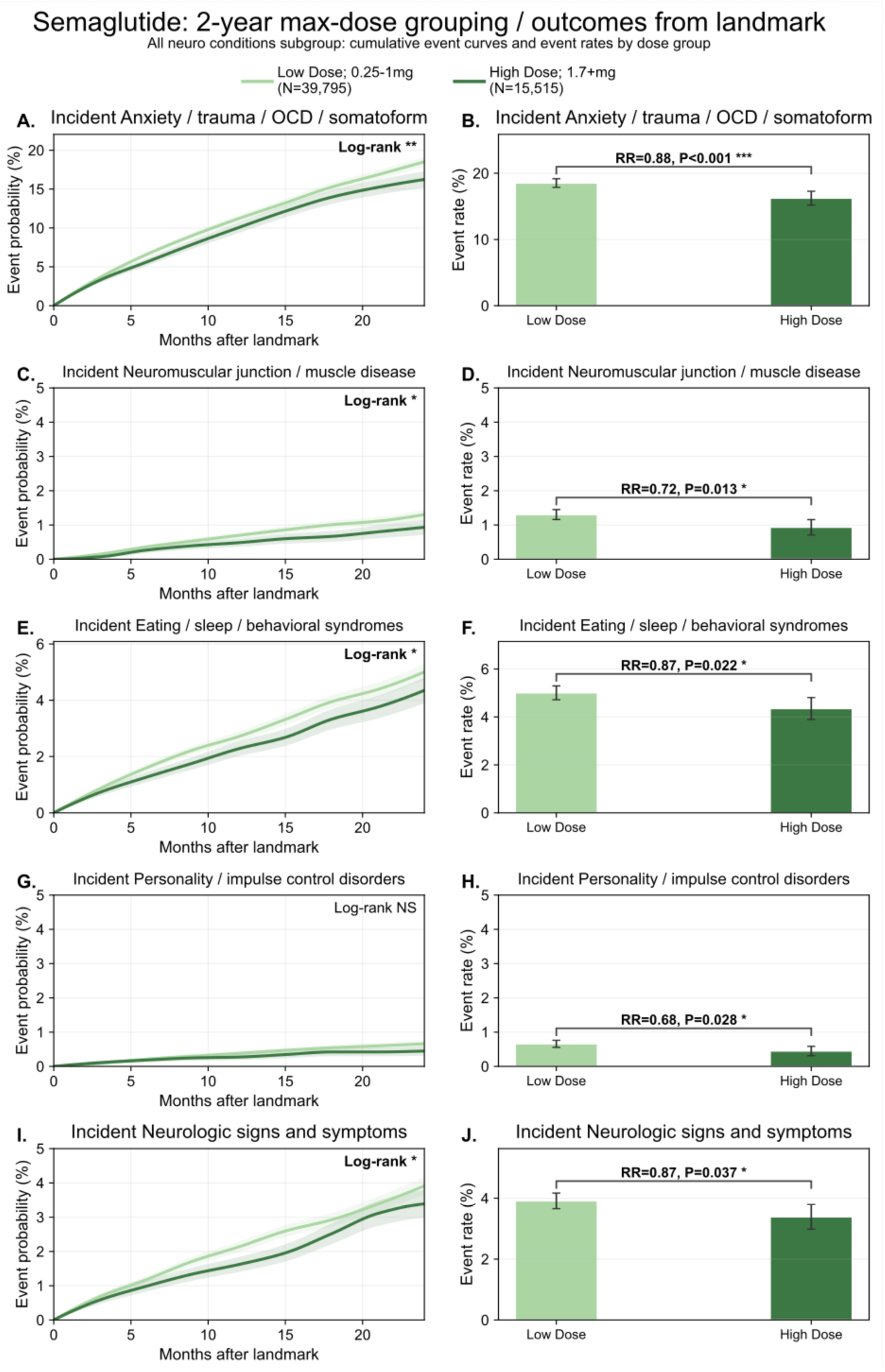
Post-landmark neurological and neuropsychiatric outcomes stratified by attained semaglutide dose during the 0–2 year landmark period. Cumulative event curves (left panels) and corresponding event-rate summaries (right panels) are shown for anxiety/trauma/OCD/somatoform disorders, neuromuscular junction or muscle disease, eating/sleep/behavioral syndromes, personality/impulse-control disorders, and neurologic signs and symptoms after landmarking at 2 years from semaglutide initiation. Patients were grouped by maximum semaglutide dose attained during the pre-landmark window as low dose (0.25–1.0 mg; N = 39,795) or high dose (≥1.7 mg; N = 15,515). Across the outcomes shown, the high-dose group had lower post-landmark event rates than the low-dose group, with significant reductions for anxiety/trauma/OCD/somatoform disorders (RR = 0.88, P < 0.001), neuromuscular junction or muscle disease (RR = 0.72, P = 0.013), eating/sleep/behavioral syndromes (RR = 0.87, P = 0.022), personality/impulse-control disorders (RR = 0.68, P = 0.028), and neurologic signs and symptoms (RR = 0.87, P = 0.037). Log-rank significance is shown within the cumulative-incidence panels, and relative rate ratios with corresponding P values are shown within the event-rate panels.

**Figure S3.**
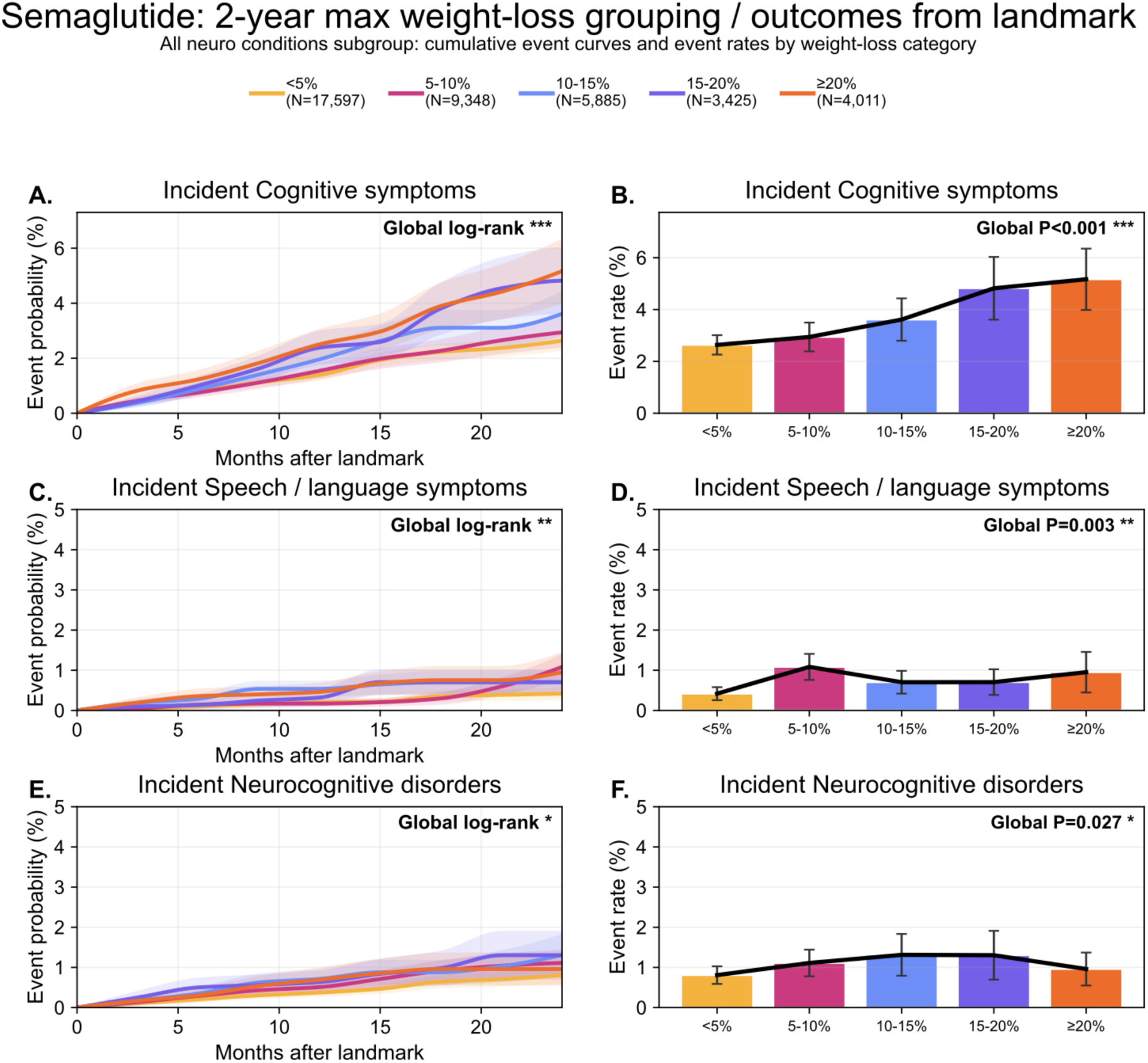
Post-landmark cognitive and language-related outcomes stratified by maximum achieved weight loss during the 0–2 year landmark period. Cumulative event curves (left panels) and corresponding event-rate summaries (right panels) are shown for cognitive symptoms, speech/language symptoms, and neurocognitive disorders across five categories of maximum body-weight change achieved before landmark: <5%, 5–10%, 10–15%, 15–20%, and ≥20%. Global log-rank statistics for the cumulative-incidence curves and global P values for the event-rate comparisons are displayed within each panel. Cognitive symptoms and speech/language symptoms showed significant variation across weight-loss strata, whereas neurocognitive disorders showed a more modest but still significant association. Overall, these analyses indicate that post-landmark cognitive and language-related outcomes were more closely aligned with pre-landmark weight-loss strata than were most other neuropsychiatric domains.

**Figure S4.**
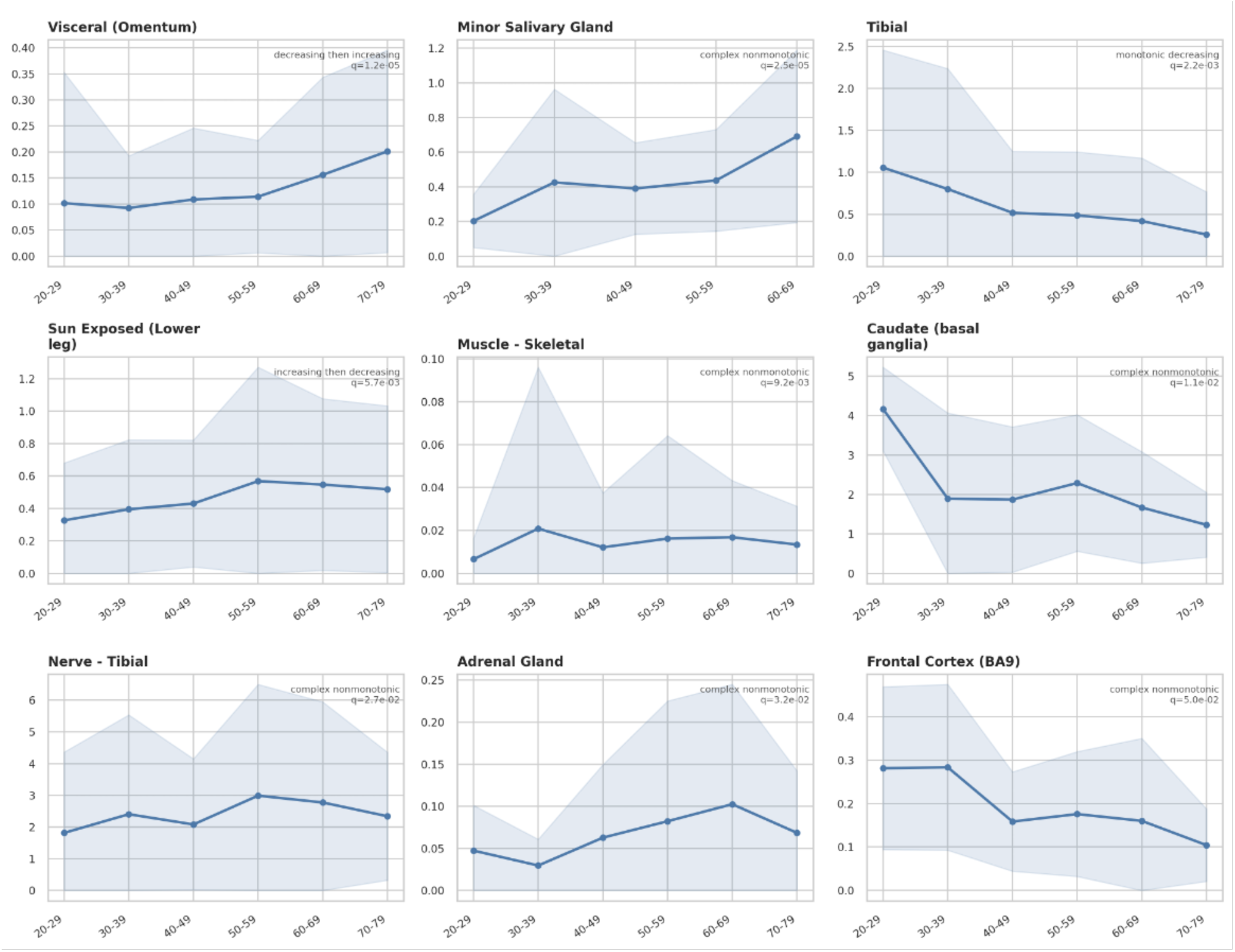
Age-stratified GLP1R expression trajectories across human tissues. Age-dependent patterns of GLP1R expression across multiple tissues are shown, stratified into decade-wise bins (20–29 through 70–79 years). Each panel represents a distinct tissue or anatomical site, with the solid line indicating mean expression and the shaded region representing variability (e.g., standard deviation or confidence interval) across samples. Distinct temporal patterns of GLP1R expression are observed across tissues. Visceral adipose (omentum) demonstrates a decreasing-then-increasing trajectory, with higher expression in older age groups. Minor salivary gland and adrenal gland exhibit complex nonmonotonic patterns, with gradual increases into mid-to-late adulthood. Tibial artery shows a monotonic decrease in GLP1R expression with age. Sun-exposed lower leg skin demonstrates an increasing-then-decreasing pattern, peaking in midlife. Skeletal muscle shows low overall expression with modest nonmonotonic variation across age groups. Neural tissues display heterogeneous dynamics: caudate (basal ganglia) and frontal cortex (BA9) exhibit declining or complex nonmonotonic trajectories, while tibial nerve shows a midlife peak followed by decline. Annotations within each panel denote the inferred trajectory class (monotonic, increasing-then-decreasing, decreasing-then-increasing, or complex nonmonotonic) along with associated statistical significance (q-values), reflecting false discovery rate–adjusted assessments of age-associated trends. These data highlight tissue-specific and nonuniform age-related remodeling of GLP1R expression, suggesting that GLP-1 receptor biology varies dynamically across the lifespan and anatomical context.

**Table S1.** Outcome definitions for neurological and neuropsychiatric condition groups using ICD-10 and ICD-9 code prefixes.

| Condition Group | Considered for outcome analysis? | ICD10 Code Prefixes | ICD9 Code Prefixes | Reason for grouping into a single outcome family |
| --- | --- | --- | --- | --- |
| **CNS infections / inflammation** | Yes | G00, G01, G02, G03, G04, G05, G06, G07, G08, G09 | 320, 321, 322, 323, 324, 325, 326 | Capture a unified class of inflammatory/infectious CNS diagnoses that are individually rare but clinically related. |
| **Viral / prion CNS infections** | Yes | A80, A81, A82, A83, A84, A85, A86, A87, A88, A89 | 045, 046, 047, 048, 049 | Considered separately from broader CNS infection codes because these conditions share distinct infectious/neurodegenerative etiologies and are too sparse to analyze reliably as standalone outcomes. |
| **Hereditary / systemic CNS atrophies** | Yes | G10, G11, G12, G13, G14 | 333, 334 | Capture progressive atrophy-linked neurodegenerative syndromes as one low-frequency but clinically coherent outcome family with shared relevance to long-latency CNS decline |
| **Parkinson's / movement disorders** | Yes | G20, G21, G22, G23, G24, G25, G26 | 332, 3330, 3331, 3332, 3334, 3335, 3336 | Clinically related movement-disorder phenotypes involving extrapyramidal or basal-ganglia dysfunction. |
| **Dementia / degenerative CNS disease** | Yes | G30, G31, G32 | 290, 3310, 3311, 3312, 3319 | Captures diagnosed degenerative cognitive disease. |
| **Demyelinating CNS disease** | Yes | G35, G36, G37 | 340, 341 | Captures multiple sclerosis and related autoimmune inflammatory disorders characterized by the destruction of myelin; grouped as a pathophysiologically unified family of neuroinflammatory conditions. |
| **Seizure and paroxysmal neurologic disorders** | Yes | G40, G41, G43, G44, G45, G46, G47 | 345, 346, 347 | Grouped to represent recurrent or episodic neurologic syndromes that are clinically event-based rather than steadily progressive, enabling analysis of an intermittently expressed neurologic burden class |
| **Cranial neuropathies / radicular syndromes** | Yes | G50, G51, G52, G53, G54, G55, G56, G57, G58, G59 | 350, 351, 352, 353, 354, 355, 356, 357 | Capture focal nerve and root syndromes across cranial and peripheral distributions. |
| **Polyneuropathies / PNS disorders** | Yes | G60, G61, G62, G63, G64, G65 | 3560, 3561, 3562, 3563, 3564, 3565, 3568, 3569 | Capture diffuse peripheral nerve disease, particularly relevant to metabolic and systemic injury. |
| **Neuromuscular junction / muscle disease** | Yes | G70, G71, G72, G73 | 358, 359 | Capture disorders affecting neuromuscular transmission and muscle integrity. |
| **Residual CNS and neurologic disorders** | Yes | G89, G90, G91, G92, G93, G94, G95, G96, G97, G98, G99 | 3490, 3491, 3492, 3498, 3499 | Residual CNS category to retain clinically relevant neurologic codes not captured by more specific predefined families. |
| **Malignant brain / CNS tumors** | Yes | C71, C72 | 191, 192 | Represents primary cancerous growths within the central nervous system, distinguishing them from benign neoplasms due to their aggressive clinical course and distinct oncology treatment pathways. |
| **Benign / uncertain CNS neoplasms** | Yes | D33, D43 | 225, 2371 | Considered separately from malignant tumors because they differ biologically and clinically. |
| **Neurocognitive disorders** | Yes | F01, F02, F03, F04, F05, F06, F07, F08, F09 | 290, 2940, 2941, 2942, 2948, 2949 | Capture formal cognitive and neuropsychiatric syndrome diagnoses. |
| **Substance-related neuropsychiatric disorders** | Yes | F10, F11, F12, F13, F14, F15, F16, F17, F18, F19 | 303, 304, 305 | Aggregates mental and behavioral disorders due to psychoactive substance use (e.g., alcohol, opioids), providing a unified family for dependencies that share common neurobiological reward-circuitry involvement. |
| **Psychotic disorders** | Yes | F20, F21, F22, F23, F24, F25, F28, F29 | 295, 297, 298 | Capture schizophrenia-spectrum and related psychotic conditions. |
| **Mood / affective disorders** | Yes | F30, F31, F32, F33, F34, F38, F39 | 296, 3004, 30113 | Capture depressive and bipolar-spectrum conditions. |
| **Anxiety / trauma / OCD / somatoform** | Yes | F40, F41, F42, F43, F44, F45, F48 | 300, 308, 309 | Capture anxiety-spectrum and stress-linked disorders that often co-occur clinically. |
| **Eating / sleep / behavioral syndromes** | Yes | F50, F51, F52, F53, F54, F55, F59 | 307 | Grouped to capture different behavioral diagnoses. |
| **Personality / impulse control disorders** | Yes | F60, F61, F62, F63, F64, F65, F66, F68, F69 | 301, 302 | Groups pervasive and persistent maladaptive patterns of behavior and inner experience (e.g., Borderline, Antisocial, Impulse Control), which are clinically distinct from episodic mood or psychotic disorders. |
| **Neurologic signs and symptoms** | Yes | R29 | 7813, 7814, 7815, 7816, 7817, 7818, 7819 | Capture nonspecific neurologic presentations that may precede formal diagnosis. |
| **Cognitive symptoms** | Yes | R41 | 78093, 78097 | Kept separate from neurocognitive disorders because these are symptom-level codes rather than formal syndrome diagnoses. |
| **Speech / language symptoms** | Yes | R47 | 7843, 7845 | Grouped as a symptom domain rather than disease-specific diagnosis |
| **Hallucinations / perceptual symptoms** | Yes | R44 | 7801 | Kept as a distinct symptom-level family because these symptoms can be seen in a number of neuropsychiatric diagnoses. |
| **Stroke / cerebrovascular disease** | No | I60, I61, I62, I63, I64, I65, I66, I67, I68, I69 | 430, 431, 432, 433, 434, 435, 436, 437, 438 | Grouped to capture acute cerebrovascular events and chronic cerebrovascular sequelae as a unified vascular-neurologic outcome domain |
| **Congenital CNS malformations** | No | Q00, Q01, Q02, Q03, Q04, Q05, Q06, Q07 | 740, 741, 742 | Grouped to capture developmental structural CNS diagnoses as a low-frequency but conceptually coherent family |
| **Intellectual disability** | No | F70, F71, F72, F73, F78, F79 | 317, 318, 319 | Grouped to capture developmental cognitive disability as a coherent neurodevelopmental category with low frequency in adult incident analyses |
| **Autism / developmental / learning disorders** | No | F80, F81, F82, F83, F84, F88, F89 | 299, 315 | Grouped to represent developmental-neurobehavioral diagnoses with overlapping childhood-onset biology and sparse adult incident counts |
| **ADHD / childhood behavioral neuropsychiatry** | No | F90, F91, F93, F94, F95, F98 | 312, 313, 314 | Grouped to capture attention, conduct, tic, and related childhood-onset behavioral syndromes as one developmental behavioral family |
| **Traumatic brain injury** | No | S06 | 850, 851, 852, 853, 854 | Distinct family because it represents an externally caused neurologic event rather than a primary endogenous neuropsychiatric disease process |
| **Cerebral palsy / paralytic syndromes** | No | G80, G81, G82, G83 | 342, 343, 344 | Grouped as chronic motor-impairment syndromes with overlapping functional consequences and low event rates when separated |

**Table S2.** Remaining neuro-outcome summaries for semaglutide dose-versus-weight-loss decomposition without any significant association to weight loss/max achieved dose.

| Outcome | N at risk | Events | Low-Dose<br>Event Rate<br>(%) | High-Dose<br>Event Rate<br>(%) | Rate ratio (High /<br>Low Dose) | P-value (High<br>vs Low<br>Dose) | P-value<br>(Weight Loss<br>Strata) |
| --- | --- | --- | --- | --- | --- | --- | --- |
| Incident Cranial neuropathies /<br>radicular syndromes | 48330 | 1852 | 7 | 6.5 | 0.922 (0.834-1.020) | 0.116 | 0.881 |
| Incident Viral / prion CNS infections | 55286 | <11 | 0 | 0 | 3.909 (0.711-21.474) | 0.117 | 0.606 |
| Incident Psychotic disorders | 55018 | 109 | 0.4 | 0.3 | 0.738 (0.489-1.115) | 0.149 | 0.743 |
| Incident Parkinson's / movement<br>disorders | 52056 | 1109 | 3.8 | 3.5 | 0.937 (0.821-1.069) | 0.335 | 0.643 |
| Incident CNS infections /<br>inflammation | 55016 | 85 | 0.3 | 0.3 | 1.231 (0.794-1.908) | 0.352 | 0.147 |
| Incident Demyelinating CNS<br>disease | 54508 | 66 | 0.2 | 0.2 | 1.268 (0.767-2.097) | 0.354 | 0.662 |
| Incident seizure and paroxysmal<br>neurologic disorders | 24220 | 3659 | 26 | 25.3 | 0.971 (0.909-1.037) | 0.374 | 0.171 |
| Incident Hallucinations / perceptual<br>symptoms | 55220 | 94 | 0.3 | 0.3 | 0.913 (0.581-1.434) | 0.693 | 0.543 |
| Incident Benign / uncertain CNS<br>neoplasms | 55010 | 49 | 0.2 | 0.2 | 1.024 (0.579-1.810) | 0.936 | 0.740 |
| Incident Dementia / degenerative<br>CNS disease | 54790 | 351 | 1.2 | 1.2 | 0.990 (0.782-1.254) | 0.937 | 0.312 |
| Incident Malignant brain / CNS<br>tumors | 55170 | 27 | 0.1 | 0.1 | 0.969 (0.438-2.142) | 0.937 | 0.451 |
| Incident Neurocognitive disorders | 54614 | 383 | 1.3 | 1.3 | 1.002 (0.804-1.249) | 0.987 | 0.050 |
| Incident Polyneuropathies / PNS<br>disorders | 51791 | 1202 | 4.1 | 4.1 | 1.000 (0.886-1.128) | 0.995 | 0.484 |

**Table S3.** Top 20 tissues by mean GLP1R TPM in the bulk RNA-seq data.

| Tissue | N samples | N subjects | Mean TPM | SD TPM | Median TPM | Min TPM | Max TPM |
| --- | --- | --- | --- | --- | --- | --- | --- |
| Pancreas - Islets | 3 | 3 | 26.367 | 6.265 | 29.353 | 19.167 | 30.580 |
| Pancreas | 362 | 362 | 6.916 | 2.482 | 6.565 | 1.465 | 16.289 |
| Pancreas - Acini | 9 | 9 | 6.509 | 2.653 | 5.936 | 3.923 | 11.971 |
| Pancreas - Mixed Cell | 10 | 10 | 5.720 | 2.164 | 5.146 | 3.415 | 9.749 |
| Heart - Atrial Appendage | 461 | 461 | 3.005 | 3.054 | 2.075 | 0.009 | 21.237 |
| Nerve - Tibial | 670 | 670 | 2.607 | 3.072 | 1.399 | 0.008 | 23.385 |
| Brain - Hypothalamus | 257 | 257 | 2.401 | 3.632 | 1.225 | 0 | 38.648 |
| Brain - Caudate (basal ganglia) | 300 | 300 | 1.903 | 1.607 | 1.519 | 0.009 | 7.707 |
| Brain - Putamen (basal ganglia) | 254 | 254 | 1.424 | 1.481 | 0.987 | 0.005 | 9.625 |
| Heart - Left Ventricle | 452 | 452 | 1.401 | 1.844 | 0.821 | 0 | 14.834 |
| Brain - Nucleus accumbens (basal ganglia) | 285 | 285 | 1.131 | 0.984 | 0.883 | 0.016 | 6.443 |
| Stomach | 407 | 407 | 1.109 | 1.270 | 0.648 | 0 | 11.168 |
| Stomach - Muscularis | 26 | 26 | 0.913 | 0.672 | 0.669 | 0.249 | 2.701 |
| Lung | 604 | 604 | 0.807 | 0.935 | 0.487 | 0 | 6.379 |
| Thyroid | 684 | 684 | 0.621 | 0.519 | 0.476 | 0.012 | 4.663 |
| Stomach - Mixed Cell | 29 | 29 | 0.565 | 0.359 | 0.512 | 0.052 | 1.529 |
| Artery - Tibial | 691 | 691 | 0.540 | 0.912 | 0.209 | 0 | 9.158 |
| Skin - Sun Exposed (Lower leg) | 754 | 754 | 0.505 | 0.559 | 0.340 | 0 | 4.516 |
| Minor Salivary Gland | 181 | 181 | 0.487 | 0.407 | 0.411 | 0 | 3.218 |
| Stomach - Mucosa | 29 | 29 | 0.448 | 0.319 | 0.398 | 0.006 | 1.286 |

**Table S4.** Significant age-associated GLP1R patterns from RNA-seq data.

| Tissue | N total | Age groups tested | Test | P | Q | Age pattern |
| --- | --- | --- | --- | --- | --- | --- |
| Adipose - Visceral (Omentum) | 587 | 6 | Kruskal_Wallis | <0.001 | <0.001 | decreasing_then_increasing |
| Minor Salivary Gland | 179 | 5 | Kruskal_Wallis | <0.001 | <0.001 | complex_nonmonotonic |
| Artery - Tibial | 691 | 6 | Kruskal_Wallis | <0.001 | 0.002 | monotonic_decreasing |
| Skin - Sun Exposed (Lower leg) | 754 | 6 | Kruskal_Wallis | <0.001 | 0.006 | increasing_then_decreasing |
| Muscle - Skeletal | 818 | 6 | Kruskal_Wallis | <0.001 | 0.009 | complex_nonmonotonic |
| Brain - Caudate (basal ganglia) | 300 | 6 | Kruskal_Wallis | 0.001 | 0.011 | complex_nonmonotonic |
| Nerve - Tibial | 670 | 6 | Kruskal_Wallis | 0.004 | 0.027 | complex_nonmonotonic |
| Adrenal Gland | 295 | 6 | Kruskal_Wallis | 0.005 | 0.032 | complex_nonmonotonic |
| Brain - Frontal Cortex (BA9) | 269 | 6 | Kruskal_Wallis | 0.008 | 0.050 | complex_nonmonotonic |

**Table S5.** Significant sex-associated GLP1R patterns.

| Tissue | Male n | Female n | Mean male | Mean female | Fold female/male | Direction | P | Q |
| --- | --- | --- | --- | --- | --- | --- | --- | --- |
| Breast - Mammary Tissue | 336 | 178 | 0.246 | 0.683 | 2.778 | female-higher | <0.001 | <0.001 |
| Skin - Sun Exposed (Lower leg) | 506 | 248 | 0.556 | 0.402 | 0.723 | male-higher | <0.001 | 0.020 |
| Lung | 414 | 190 | 0.907 | 0.587 | 0.647 | male-higher | 0.002 | 0.035 |
| Stomach - Muscularis | 18 | 8 | 0.740 | 1.303 | 1.762 | female-higher | 0.003 | 0.040 |
| Pituitary | 229 | 84 | 0.092 | 0.162 | 1.761 | female-higher | 0.004 | 0.040 |

